# Incorporating phenotype heterogeneity in disease GWAS improves power while maintaining specificity

**DOI:** 10.64898/2026.03.26.26349370

**Authors:** Jasper Hof, Chao Ning, Liam Quinn, Doug Speed

## Abstract

Common complex diseases are clinically heterogeneous, yet most genome-wide association studies (GWAS) assume cases are genetically homogeneous. This challenge is compounded in large-scale biobanks, which increasingly combine cases ascertained under different recruitment strategies, raising concerns that heterogeneous case definitions may dilute genetic signal. To address this, we developed StratGWAS, a scalable framework that leverages clinical features of heterogeneity to construct a transformed phenotype that better reflects genetic liability within diseases. StratGWAS stratifies cases using secondary phenotypic information such as age of onset, medication burden, or recruitment definition. StratGWAS then estimates genetic covariance between strata, and derives a transformed phenotype that upweights cases with higher inferred genetic liability. Through simulation studies (*N* = 100k) and application to the UK Biobank (*N* = 368k), we show that StratGWAS consistently outperformed standard GWAS methods. Applied to 21 UK Biobank traits, StratGWAS upweighted individuals with earlier disease onset and higher medication burden, yielding respectively 17% and 4% more independent genome-wide significant loci than standard case–control GWAS. Applied to depression, StratGWAS upweighted individuals with multiple diagnoses, greater psychiatric comorbidity, or higher self-reported depressive symptoms, identifying eight additional independent loci compared to case-control GWAS.

## Introduction

Common complex diseases are clinically heterogeneous, differing in symptom presentation, disease severity, progression, and/or treatment response (1). This has motivated substantial interest in identifying features that distinguish cases into more homogeneous disease subgroups, enabling improved characterization of biological mechanisms and more precise risk stratification (2-8). Growing evidence indicates that clinically defined subgroups also capture differences in underlying genetic liability (9, 10), as demonstrated for major depression (6, 11, 12) and autism (13).

These differences in genetic liability are further compounded by the increasingly diverse recruitment strategies used in large-scale data collection: cases ascertained through clinical settings, self-report, or electronic health records may differ substantially in disease severity and genetic burden (14, 15), yet are routinely combined without accounting for these differences. Combining shallow and deep phenotyping definitions may dilute genetic signal and complicate interpretation, and it remains debated whether such cases should be analyzed jointly at all (16-18). Despite these challenges, most genome-wide association studies (GWAS) analyze all cases jointly in a binary case-control framework, often ignoring heterogeneity in genetic liability across subgroups and potentially limiting statistical power (19, 20).

Here we introduce StratGWAS, a statistical framework that explicitly models heterogeneity in genetic liability across disease subgroups. StratGWAS stratifies cases based on user-defined variables and infers subgroup-specific genetic liability from the data, yielding a transformed phenotype that more closely approximates underlying genetic liability and increases detection power for association testing, while maintaining disease specificity.

Using simulations and data from the UK Biobank (21), we show that incorporating clinical measures for heterogeneity can substantially improve locus discovery. Across 21 common diseases stratified by age at onset and medication burden, StratGWAS reveals distinct heritability estimates across subgroups and identifies additional trait-associated loci. When applied to major depressive disorder (MDD), StratGWAS identified distinct disease characteristics associated with higher genetic liability and discovers additional loci associated with MDD. Importantly, StratGWAS maintained disease specificity, showing replication rates similar to traditional GWAS and achieving similar genetic correlations. Together, these results demonstrate that leveraging routinely collected clinical information can substantially enhance genetic discovery in complex disease.

## Results

### Overview of StratGWAS

A detailed description of StratGWAS is included in the Methods and **Supplementary Notes 1-2**. In brief, StratGWAS stratifies cases into subgroups based on user-defined variables, such as age at onset, medication burden, symptom severity, or recruitment source (**Figure 1**). We refer to these variables as “stratification variables”, which can be either continuous or categorical. Next, StratGWAS estimates subgroup-specific heritabilities and genetic covariances using an approach analogous to SumHer (22), then assembles these estimates into a genetic covariance matrix. StratGWAS computes subgroup-specific weights from the genetic covariance matrix, that estimate the average genetic liability in each disease subgroup. Based on these weightings, each individual is assigned a new “transformed phenotype”, which better approximates genetic liability and maximizes heritability as a function of subgroup membership. The transformed phenotype is used as input for GWAS analysis to identify SNPs associated with target binary trait.

**Fig. 1.**
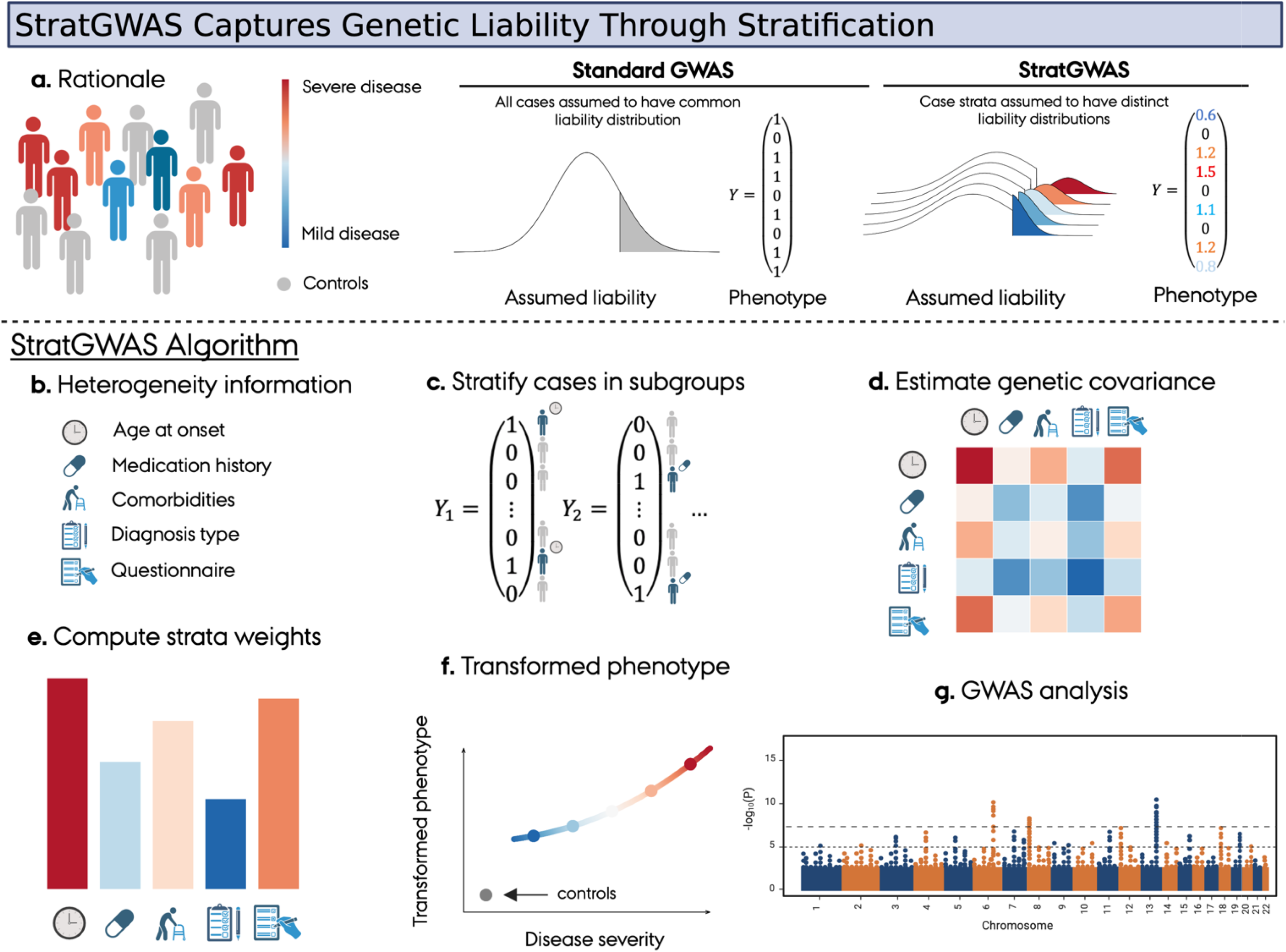
Rationale and algorithmic design of StratGWAS. StratGWAS treats clinical heterogeneity among cases as a proxy for differences in genetic liability, constructing a transformed phenotype that improves GWAS power. **a**. Common complex diseases are clinically heterogeneous, with cases differing in disease severity and genetic liability. Standard case-control GWAS treat all cases equivalently, failing to leverage this information. **b**. Examples of clinical variables that may index differences in genetic liability among cases. **c**. StratGWAS divides cases into subgroups based on user-defined stratification variables, constructing separate binary phenotypes for each subgroup against a shared set of controls coded as 0. **d**. Subgroup-specific heritabilities and genetic covariances are estimated from summary statistics and assembled into a genetic covariance matrix summarizing the shared genetic architecture across strata. **e**. Each stratum is assigned a weight derived from the leading eigenvector of the genetic covariance matrix. **f**. These weights are used to construct individual-level transformed phenotypes by scaling each individual’s phenotype according to the estimated genetic liability of their stratum. **g**. StratGWAS performs a GWAS using the transformed phenotype.

### Data

We used genotype and phenotype data from the UK Biobank (21) (application number: 21432), comprising 487,152 individuals, of whom approximately 95% are of European ancestry. Our main dataset contains 409k white British individuals, from which we randomly sampled a set containing 90% of individuals (368k participants) for GWAS analyses. For simulations, we also use a subset of 100,000 unrelated white British individuals, selected so that no pair of individuals had a closer relationship than that of second cousins. After restricting to autosomal, biallelic, directly genotyped SNPs with MAF > 0.001, the genotype data contained 690,264 SNPs, while we additionally restricted to SNPs with MAF > 0.01 for the simulation dataset, yielding 641,204 SNPs. Detailed descriptions of data access, quality control procedures, and cohort-specific preprocessing steps are provided in the **Supplementary Note 3**.

For analysis of real traits, we selected 21 binary traits based on three criteria: (i) availability as 3-digit ICD-10 codes in the UK Biobank; (ii) prevalence greater than 5%; and (iii) observed-scale heritability greater than 0.05, as estimated using SumHer (22). All traits were analyzed using StratGWAS based on two stratification variables: (i) disease age at onset and (ii) medication burden within disease category. Medication burden was defined as the number of unique medications prescribed to an individual, where we required that the medications were from a British National Formulary (BNF) category that matched the disease based on its ICD-10 Chapter (see **Supplemental Note 3**). The resulting transformed phenotypes were analyzed using both linear regression and the mixed-model association analysis tool LDAK-KVIK (23).

We additionally analyzed major depressive disorder, defined here as broad depression (24) based on self-reported help-seeking behavior for mental health difficulties from a general practitioner or psychiatrist (UK Biobank field codes 2090 and 2100). We used ten stratification variables: (i) source of depression diagnosis (general practitioner, psychiatrist, or both); (ii) diagnosed with F32; (iii) diagnosed with F33; (iv) offered or sought treatment for depression; (v) diagnosed with bipolar disorder; (vi) diagnosed with schizophrenia; and self-reported frequency of (vii) depressed mood; (viii) disinterest; (xi) restlessness; and (x) tiredness. GWAS results from the StratGWAS-derived phenotype were compared with those from a standard case-control GWAS, and validated using and a recent MDD GWAS from the Psychiatric Genetics Consortium (PGC) (25) that excluded the UK Biobank. Details on stratification variables are included in **Supplementary Note 3**. All analyses included the first ten principal component as covariates, alongside age, age^2^, sex, and age × sex.

### Comparison with existing methods

For our real data application based on age at onset and medication burden, we additionally compared StratGWAS with GenomicSEM (26) and ADuLT (27). For our implementation of GenomicSEM, we used the subgroup-specific summary statistics obtained from StratGWAS to perform a multivariate GWAS assuming one common factor. The application of ADuLT was restricted to the analyses based on age at onset, with liabilities estimated using cumulative incidence proportions based on birth year strata of five years, stratified by sex. With regard to family history in ADuLT, we included information of first-degree relatives which we inferred using KING software (28). More details on the implementation of GenomicSEM and ADuLT are provided in the **Supplementary Note 4**.

### StratGWAS controls type 1 error

We evaluated StratGWAS using simulated binary traits for 100,000 individuals across a range of trait prevalences, heritabilities, and stratification variable configurations (**Figure 2**). Specifically, we simulated combinations of target binary trait and stratification variables with different degrees of shared liability and genetic correlation, by simulating SNPs that were either causal for both the target binary trait and the stratification variable, or causal for the stratification variable only.

**Fig. 2.**
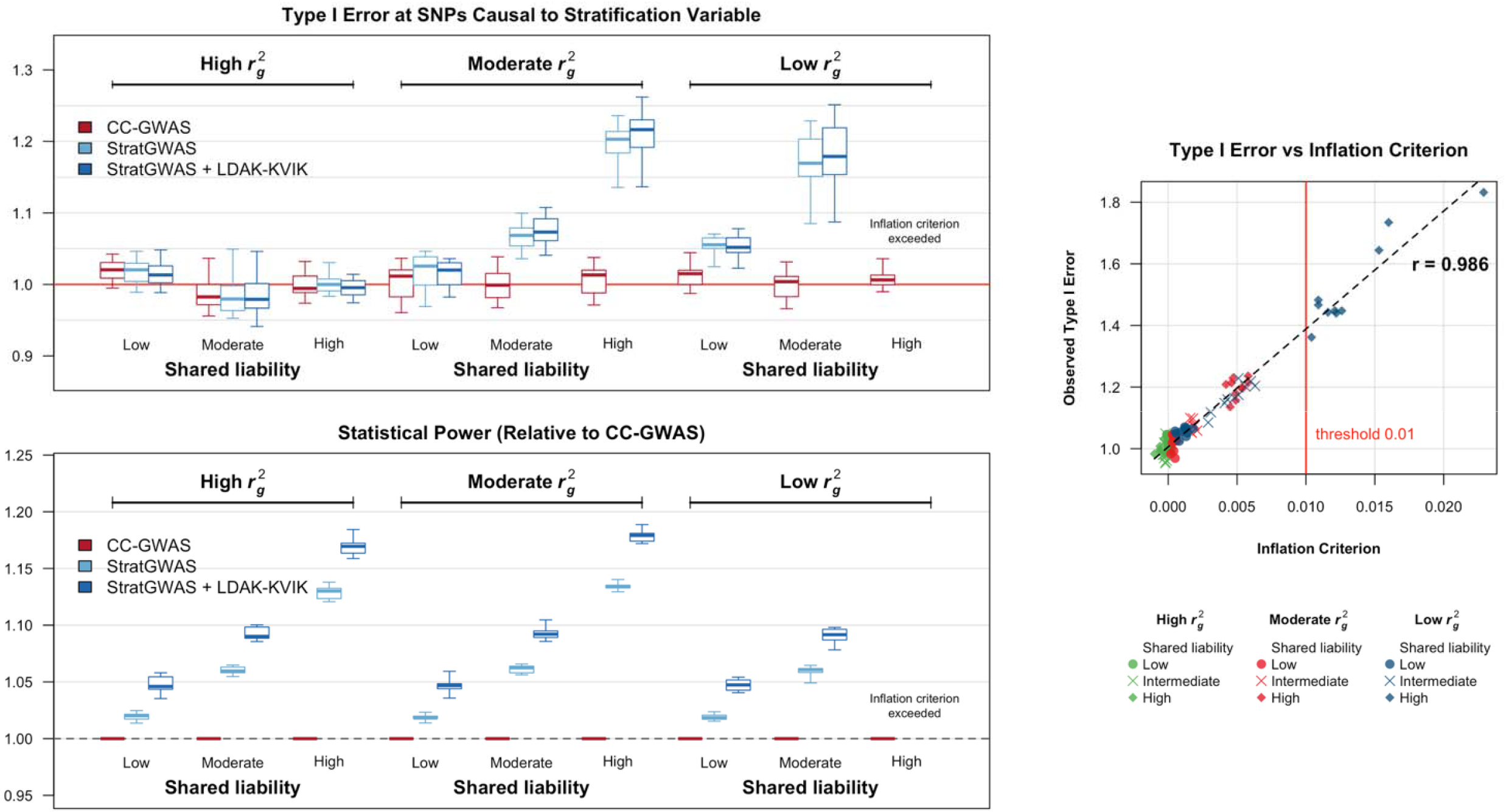
Statistical power and type 1 error of StratGWAS versus case-control GWAS across different scenarios of stratification variables. We generated 10 binary traits for 100,000 individuals with prevalence 0.5 and liability heritability 0.2. For each phenotype, we generated nine associated stratification variables that varied in shared liability and genetic correlation with the target trait by introducing genetic effects specific to stratification variable. Panels report the mean chi-squared test statistic of causal SNPs relative to case-control GWAS (top left), and mean chi-squared test statistic of SNPs that are causal to the stratification variable, but not the binary trait (bottom left). The right panel plots the estimated inflation criterion against observed type 1 errors for all simulated combinations, with the fitted correlation shown in the top right corner. The red vertical line indicates the default inflation criterion threshold; StratGWAS results are not shown for the scenario exceeding this threshold (low and high shared liability), as we do not recommend using StratGWAS in these situations.

We first evaluated type 1 error control at SNPs causal for the stratification variable, but not for to the target trait. Here we observed increased risk of false positives from analyzing the transformed phenotype whenever the stratification variable had imperfect genetic correlation to the target binary trait. To address this, StratGWAS computes an “inflation criterion”, which quantifies the risk of genetic confounding introduced by the stratification variable. Across all simulation scenarios, the inflation criterion showed high correlation to observed type 1 error rates (**Figure 2, Supplementary Figure 1**). In one of nine simulation scenarios considered, the criterion exceeded 0.01, which is the default threshold at which StratGWAS decides a stratification variable should be excluded. Across the eight scenarios where the stratification variable was retained, none of the null SNPs that were causal for stratification variable reached genome-wide significance. In contrast, among phenotypes excluded by the inflation criterion, 14 null SNPs were genome-wide significant. We also examined type 1 error at SNPs not causal for either the stratification variable or the target binary trait. Here, all methods had well-controlled type 1 error (**Supplementary Figure 2**).

### StratGWAS increases statistical power in simulations

Across all simulation scenarios considered, StratGWAS demonstrated consistently higher statistical power than case-control GWAS, as measured by the mean chi-squared statistic at causal SNPs, except for the one scenario where the inflation criterion exceeded the 0.01 threshold (where StratGWAS ignores the stratification variable and is thus equivalent to a regular case-control GWAS) (**Figure 2**). Power increased with the degree of shared liability between the stratification variable and target trait. The largest gains were observed for traits with higher prevalence and heritability (**Supplementary Figure** 1), likely reflecting more precise heritability estimates within strata. Power was further improved when the StratGWAS-transformed phenotype was analyzed using LDAK-KVIK (23).

The transformation weights estimated by StratGWAS closely reflected the mean genetic liability of each subgroup (**Supplementary Figure 3**). Discrimination between subgroups with low and high genetic liability improved as the shared liability between binary and stratification variable increased.

Conversely, when genetic correlation was low, subgroup weights converged, reflecting reduced genetic information in the stratification variable.

### Upweighting earlier disease onset improves power

When applied to 21 binary traits in the UK Biobank, StratGWAS consistently upweighted individuals with earlier disease onset: across traits, individuals in the earliest onset quintile received weights on average 2.16-fold higher than those of the latest quintile (**Figure 3, Supplementary Figure 4, Supplementary Tables 1-2**). This translated into a 17% increase in independent genome-wide significant loci relative to case-control GWAS. Combining the StratGWAS transformed phenotype with LDAK-KVIK further improved discovery to 20% more loci, compared with an increase of 14% for ADuLT and 4% decrease for GenomicSEM.

**Fig. 3.**
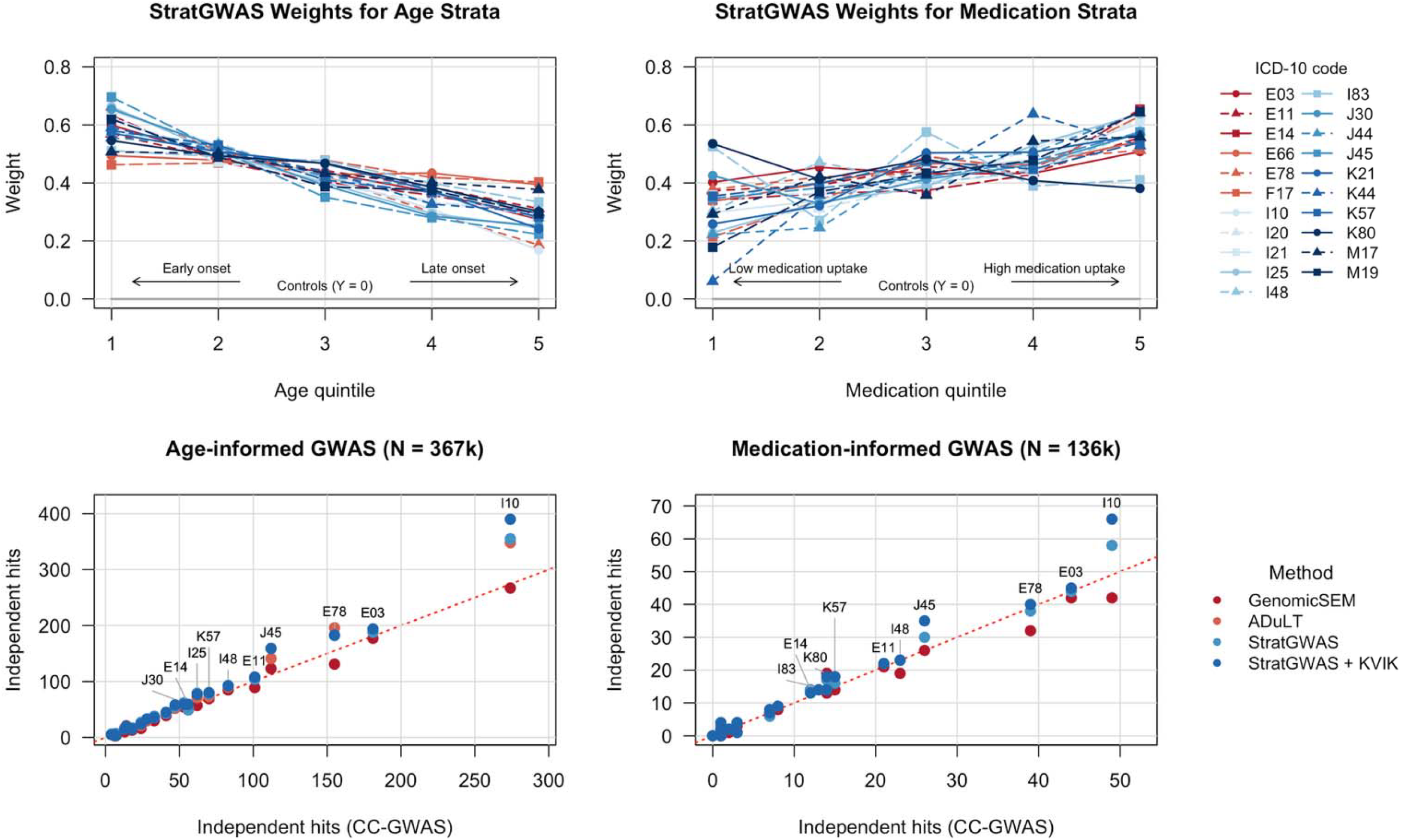
Application and comparison of StratGWAS to 21 common binary traits in the UK Biobank based on age at diagnosis and medication burden. We analyzed 21 binary traits defined by ICD-10 codes in the UK Biobank, using age at onset (left panels) and medication burden (right panels) as stratification variables. The top panels show the StratGWAS weights assigned to each quintile-based stratum, reflecting the relative genetic liability of each subgroup. The bottom panels compare the number of independent genome-wide significant loci from the GWAS tools considered with case-control GWAS. The diagonal red dotted lines represent the case where the number of independent hits matches that of case-control GWAS.

The ranking of methods was similar when considering the number of replicated loci in FinnGen (29) (**Supplemental Table 3**). Genetic correlations between StratGWAS summary statistics and FinnGen were comparable to those from standard GWAS (mean = 0.83 for case-control GWAS, mean = 0.82 for StratGWAS), suggesting that the transformed phenotype preserves the specificity of the binary trait (**Supplementary Table 4**).

### StratGWAS upweights individuals with higher medication burden

We next constructed a stratification variable reflecting medication burden using primary care prescription data from 180,026 UK Biobank participants, which we defined as the number of unique medication types within the same BNF chapter as binary disease, divided by age (**Supplementary Table 5**). Cases in the top quintile (i.e., those in the 20% with highest medication burden) received on average 1.65-fold higher weights than those in the bottom quintile. The StratGWAS transformed phenotype yielded 4% more independent genome-wide significant loci than case-control GWAS, and 11% more than GenomicSEM (**Figure 3, Supplementary Figure 4**). The more modest improvement relative to age-at-onset analyses likely reflects weaker shared genetic liability between medication burden and disease status and may be addressable through more disease-specific measurements of treatment intensity. Replication rates in FinnGen and genetic correlations with FinnGen summary statistics were consistent to those from case-control GWAS, supporting the validity of the transformed phenotypes (**Supplementary Tables 3–4**).

### StratGWAS identifies more depression loci by upweighting severe cases

When applied to major depressive disorder (MDD), StratGWAS assigned higher transformed phenotypes to individuals who had sought help both from a doctor and psychiatrist, those diagnosed with bipolar and/or schizophrenia, and who had higher self-reported depression-related symptoms, such as tiredness and disinterest (**Figure 4a, Supplementary Figures 5-6, Supplementary Table 6**). For comparison, **Figure 4b** reports the marginal weights assigned to each category (i.e., the impact on the transformed phenotype of a category when all other categories are kept fixed). It is interesting to note that the marginal weights for bipolar and schizophrenia are negative; therefore, the fact these individuals tend to higher transformed phenotypes in **Figure 4a** must reflect the high correlations between categories (for example, we note that people with schizophrenia are more likely to report high levels of restlessness), and that the questionnaire-based categories are assigned larger weights than the comorbidity categories.

**Fig. 4.**
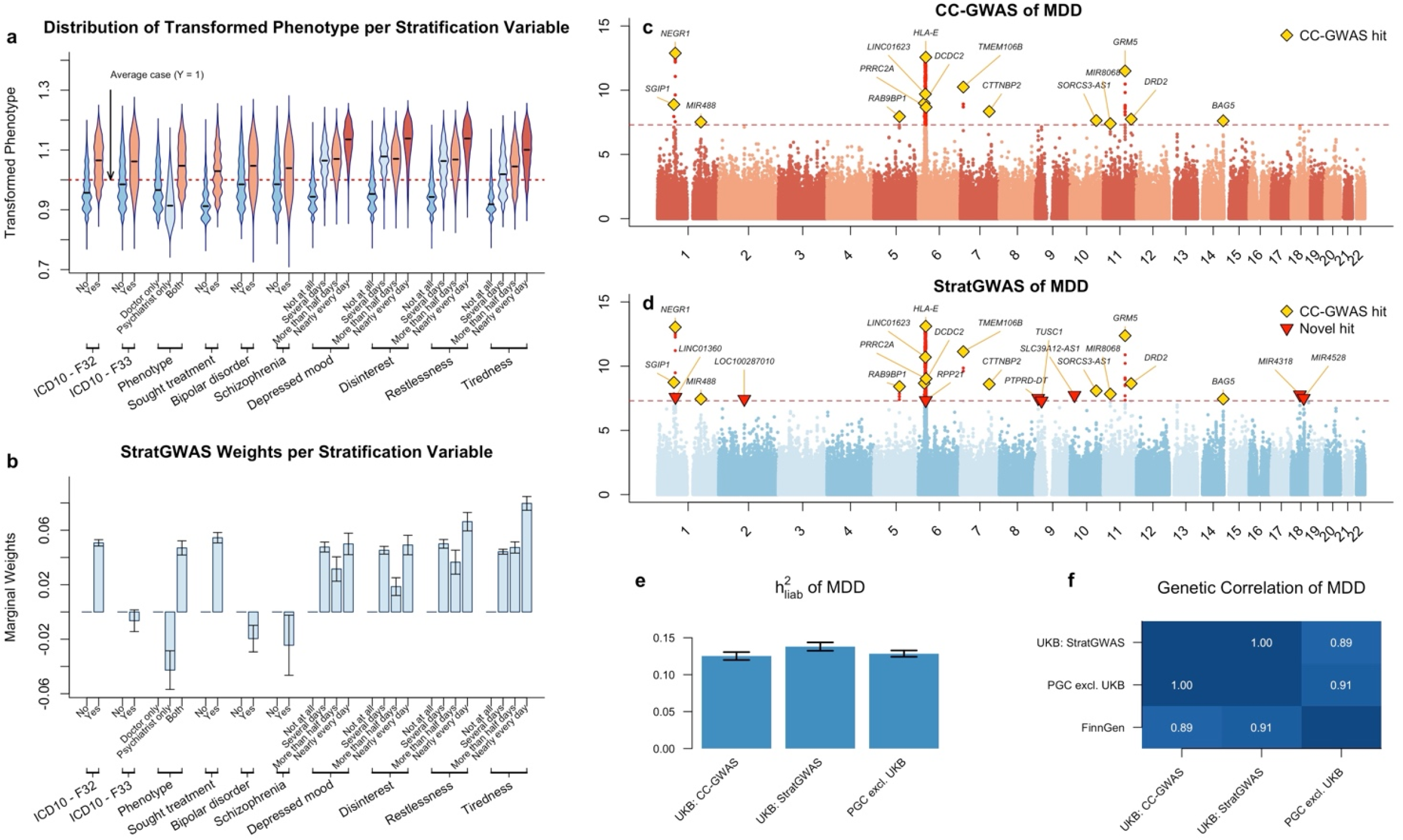
Application of StratGWAS to major depressive disorder in the UK Biobank. **a**. Violin plots showing the distribution of the StratGWAS transformed phenotype across subgroups of each stratification variable. **b**. Bar plot of StratGWAS marginal weights with standard errors for all categorical variables within each stratification variable. Weights are standardized such that for each stratification variable, the first category serves as reference. **c, d**. Manhattan plots for case-control GWAS and StratGWAS of MDD. **e, f**. Heritability and genetic correlation estimates of MDD derived from case-control GWAS, StratGWAS, and PGC (excluding UK Biobank) summary statistics, computed using SumHer (22).

The StratGWAS-derived phenotype identified 23 independent significant loci compared to 15 from case-control GWAS, all of which were also significant in StratGWAS (**Figure 4c-d**). Validation analyses supported the suitability of the transformed phenotype: it achieved a genetic correlation of 0.91 with PGC MDD summary statistics, compared with 0.89 for case-control GWAS, while seven of the eight SNPs detected by StratGWAS but missed by case-control GWAS were replicated in the PGC summary statistics (P < 0.05/23).

### StratGWAS is computationally efficient

StratGWAS requires approximately 1 CPU hour to analyze 100,000 individuals for a continuous stratification variable with 5 subgroups, and 5 CPU hours when analyzing 367,000 individuals (**Supplementary Table 7**). Runtime increases when the number of strata increases, but can be reduced when using multiple threads. StratGWAS has a low memory footprint: when using the default block size of 1,000 SNPs, it requires 6GB memory to analyze 100,000k individuals, and 20GB to analyze 368,000 individuals. StratGWAS supports parallel computing, which achieves approximately two-fold reduction in runtime when analyzing a single quantitative stratification variable using five threads.

### Alternative strategies for leveraging secondary phenotypes

In an earlier implementation, we jointly modelled the target binary trait and stratification variable using a linear combination of *Y, S*, and *S*^2^ (where *Y* and *S* denote the binary trait and stratification variable, respectively), deriving a transformation that maximizes heritability (**Supplemental Note 5**). While this approach models genetic liability more directly as a function of the stratification variable, it produced high levels of false-positive associations when *Y* and *S* only shared moderate genetic liability. We found that the stratification-based design of StratGWAS is more robust in these settings, as it leverages subgroup-specific genetic covariance estimated from the target binary trait alone.

### Considerations when using StratGWAS

The performance of StratGWAS depends on the accuracy of heritability and genetic covariance estimates. When stratum sample sizes are small, imprecise estimates can yield noisy transformation weights and reduced power. We therefore recommend that each stratum contains at least 1,000 cases. Although StratGWAS is robust to uninformative stratification variables, it can suffer from reduced power when many genetically irrelevant traits are included as stratification variables, as they dilute the transformed phenotype. We therefore recommend selecting stratification variables that plausibly reflect disease severity or clinically meaningful subtypes. Finally, for computational efficiency and accurate heritability estimation, we recommend using 500,000 to 1 million common SNPs to run StratGWAS.

## Discussion

We have presented StratGWAS, a framework for integrating clinical heterogeneity into GWAS by constructing a transformed phenotype that better reflects underlying genetic liability of disease. The key motivation behind StratGWAS is that case-control GWAS discards clinically meaningful variation among cases that is partly genetic in origin. For example, individuals who develop disease earlier, or are ascertained through deep phenotyping, likely carry a higher concentration of disease-predisposing variants. StratGWAS recovers this signal by stratifying cases into subgroups and assigns higher weights to those conferring a higher genetic burden. The results of our study suggest that this approach improves statistical power while maintaining disease specificity, thereby addressing current challenges in heterogeneity and phenotyping in large-scale biobank analyses.

Simulation results clarify under which conditions StratGWAS is most effective. Power gains were greatest when the stratification variable shared substantial genetic liability with the target trait, and for traits with higher prevalence and heritability where subgroup covariance can be estimated precisely. These findings suggest that variables that plausibly reflect disease severity will yield the greatest benefit, while genetically uninformative variables contribute little.

When applied to age-at-onset across 21 UK Biobank traits, StratGWAS yielded 17% more independent loci than case-control GWAS, increasing to 20% when combined with LDAK-KVIK. The consistent upweighting of individuals of earlier-onset cases is in line with prior evidence that individuals who develop disease at younger ages tend to confer greater genetic risk (30, 31). Gains from stratification by medication burden were more modest (4%), likely reflecting the broad BNF chapter-level definition used here; more disease-specific measures of treatment intensity may yield larger improvements.

In MDD, StratGWAS upweighted individuals with multiple modes of diagnosis, psychiatric comorbidities, and higher self-reported symptom burden, identifying 23 independent loci compared to 15 from case-control analysis. For this application, we found that mood-related questionnaire items were particularly informative for the transformed phenotype, which is consistent with prior evidence of elevated heritability in symptom-defined MDD subgroups (32), and suggests that symptom measures may identify subgroups carrying greater genetic susceptibility to disease. The high genetic correlation between the StratGWAS-derived phenotype and PGC MDD summary statistics (*r*_*g*_ = 0.91 versus *r*_*g*_ = 0.89 for case-control GWAS) suggests that the transformed phenotype maintains disease specificity while improving statistical power.

A key safeguard in StratGWAS is its inflation criterion, which identifies continuous stratification variables at risk of inducing false positive associations. This provides a key methodological contribution for machine-learning based strategies for inferring disease liability, which are fundamentally at risk of false positives by leveraging variables that can confound signals. We note that the inflation criterion can not be viewed as an exact measurement for the expected number of false positives, as this also depends on other factors such as sample size. Nonetheless, we recommend using this inflation criterion as a standard check to safeguard against inflation from continuous stratification variables.

StratGWAS is particularly relevant in the context of large-scale biobanks, which increasingly combine cases ascertained through self-report, primary care records, hospital episodes, and specialist assessment. These sources can differ substantially in phenotype depth, and consequently in the average genetic liability of ascertained cases (15, 33). Rather than treating this heterogeneity as a confound to be removed through strict case definitions at the cost of sample size, StratGWAS tried to exploit differences in signals among case subgroups. Using ascertainment source as a stratification variable allows broadly and deeply phenotyped cases to be analyzed jointly, with weights reflected inferred genetic liability rather than diagnostic signals (34). This offers a principled solution to outstanding concerns about combining cases from heterogeneous recruitment pipelines (16), and may be especially valuable for psychiatric traits where case definitions vary substantially across cohorts (12).

Several limitations deserve consideration. First, StratGWAS relies on accurate estimation of genetic covariance between subgroups, which requires adequate within-stratum sample sizes and may be unreliable for diseases with few cases. Second, the current framework assumes a linear relationship between subgroup liability and genetic signal, while allowing for interactions among subgroups could potentially improve performance in settings with more complex heterogeneity structure. Third, StratGWAS assumes that disease subgroups share a common liability scale and may therefore be less suitable when subgroups differ substantially in genetic architecture. Fourth, StratGWAS can produce false positives when there is a high degree of related individuals. To mitigate this, we recommend using the transformed phenotype in LDAK-KVIK (23). Finally, while we find that the inflation criterion provides robust protection against false positives in the simulation scenarios considered, its performance in the presence of environmental correlations between stratification variable and target trait, or structured populations, merits further investigation.

Overall, the utility of phenotypic stratification in StratGWAS is likely to grow as biobanks expand their depth of clinical characterization. Longitudinal disease trajectories, electronic health record-derived severity scores, and multimodal clinical assessments all represent potential stratification variables that may better capture variation in genetic liability than binary case definitions. More broadly, StratGWAS illustrates that targeted use of clinical heterogeneity can be used to define phenotypes that more closely reflect underlying genetic liability, improving GWAS power while maintaining disease specificity.

## Data availability

Our study used data from the UK Biobank, which we applied for and downloaded from www.ukbiobank.ac.dk. The UK Biobank has ethics approval from the North West Multi-centre Research Ethics Committee.

## Code availability

StratGWAS is available as R package, which can be downloaded from https://github.com/JasperHof/StratGWAS.

## Acknowledgements

This research was conducted using the UK Biobank Resource under application number 21432. The computing for this project was performed on the GenomeDK cluster (Aarhus University). We thank A. Halager and D. Søndergaard (Aarhus University) for programming suggestions, F. Tabrizi (Mid Sweden University) for assistance with phenotyping, and E. Pedersen (Aarhus University) for assistance with analysis using ADuLT. D.S. is supported by a European Research Council Consolidator Grant (ID 101088901, acronym ClassifyDiseases).

## Author contributions

J.H. and D.S. jointly developed the software, J.H. performed the analysis, J.H., C.N., L.Q. and D.S. wrote the manuscript.

## Methods

### StratGWAS framework

We outline the key features of StratGWAS below, with further details in **Supplementary Note 2**.

StratGWAS divides cases into *K* subgroups based on user-specified stratification variables, which may be continuous or categorical. For continuous variables, cases are ranked and split into *K* equally sized strata. By default, *K* = 5, such that cases in the lowest 20% are assigned to group 1, those in the 20%-40% range to group 2, etc. For categorical variables, cases are assigned to subgroups corresponding to each categorical outcome. For each case subgroup *k*, a separate phenotype *Y*_*k*_ is constructed, where controls are coded as 0 and cases within subgroup *k* are coded as 1. Linear regression is then performed independently for each *Y*_*k*_, yielding subgroup-specific summary statistics.

StratGWAS estimates subgroup-specific heritability 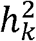 and the pairwise genetic covariance between Subgroups *γ* _*i,j*_ from summary statistics, using an approach analogous to SumHer (22) (details in **Supplemental Note 2**). These estimates are assembled into a *K* × *K* matrix Ψ

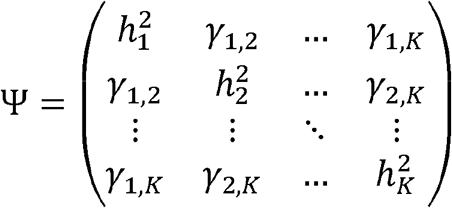

StratGWAS then computes an eigendecomposition of Ψ such that Ψ = *LDL*^⊤^, where *L* = (*L*_*1*_,…, *L*_*k*_) contains the eigenvectors and *D* is the diagonal matrix. By construction, each eigenvector is orthogonal to all previous eigenvectors and maximizes the variance of the corresponding linear combination of subgroup phenotypes. In particular, the first eigenvector *L*_1_ = (*l*_1_,…, *l*_*k*_)^⊤^ defines the linear combination of subgroups that maximizes overall heritability.

The transformed phenotype*Y*^*′;*^ constructed by StratGWAS for subsequent GWAS analysis is defined as

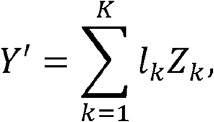

where*Z*_*k*_ represents the*n* -dimensinal indicator equal to 1 for cases in subgroup*k*, and 0 otherwise. Controls are thus assigned a value of 0 in the transformed phenotype, while cases in subgroup*k* receive weight*l*_*k*_.

### Stratification by a continuous covariate

When strata are constructed from a continuous variable, such as age at onset, or medication burden, the transformed phenotype*Y*^′;^ can be refined using smoothing, enabling it to more closely reflect the underlying continuous relationship between the stratification variable and genetic liability, and improving statistical power (**Supplemental Figure 3**).

Suppose the phenotype*Y* is divided into*K* strata based on the break points*t*_(1)_,*t*_(2)_, …,*t*_(*k*−1)_ such that stratum*k* comprises cases for whom the stratification variable*s* satisfies*t*_(*k*−1)_ ≤*S* <*t*_(*k*)_, with*t*_(0)_ = − ∞ and*t*_(*K*)_ = ∞. Further, let*m*_*k*_ denote the median stratification variable within each stratum. Having estimated*l*_*k*_, StratGWAS fits a cubic spline 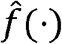 through the pairs (*m*_1_,*l*_1_), …, (*m*_K_,*l*_K_). The smoothed transformed phenotype is then defined as 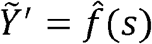 for cases and 0 for controls.

### Using multiple input stratification variables

StratGWAS can incorporate multiple stratification variables. Suppose*M* input stratification variables are provided, yielding*K*_1_, …*K*_*M*_ subgroups respectively. StratGWAS jointly considers all*K* =*K*_1_+…*K*_*M*_ subgroups, constructing a corresponding set of subgroup-specific phenotypes and estimating the genetic covariance matrix across all subgroups. An eigendecomposition is then performed to obtain subgroup weights, analogous to the single-variable case. The final transformed phenotype is constructed by combining all weighted subgroups into a single phenotype.

### Integration with LDAK-KVIK

We recommend analyzing the transformed phenotype obtained from StratGWAS using the mixed-model association analysis tool LDAK-KVIK (23). This approach further improves statistical power by modelling effects of SNPs distal to the SNP being tested and enables control of false positives arising from population structure. It also facilitates the use of a saddlepoint approximation (SPA), which ensures robust P-values when testing SNPs with low minor allele frequency (MAF) or traits with severe case-control imbalance. For our simulations and real data application, we present both the results obtained from directly analyzing the StratGWAS phenotype, as well as integrating the StratGWAS phenotype with LDAK-KVIK.

### Estimating confounding due to stratification variable

When the transformed phenotype*Y* ^′;^ is constructed by stratifying on a continuous covariate with imperfect genetic correlation to the target binary trait, there is a risk of false positive associations. We quantify this risk by modeling the liability for the binary trait and continuous stratification variable*S* as

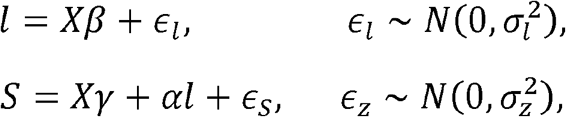

where*X* denotes the genotype matrix,*β* denotes SNP effects of the target trait,*γ* denote SNP effects specific to*S, ϵ*_*l*_ and*ϵ*_*S*_ are independently residual terms, and*α* captures shared liability. We observe the binary phenotype*Y* = 1_*l>τ*_ where liability threshold*τ* is determined by trait prevalence.

Substituting the expression of*l* into*S* yields*S* =*X (γ + α β*) +*α ϵ*_*l*_ +*ϵ*_*S*_; with genetic component*X (γ + α β*) . Defining 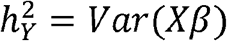 and 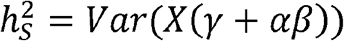 as the liability-scale heritabilities of*Y* and*S* respectively, the genetic correlation between*Y* and*S* is

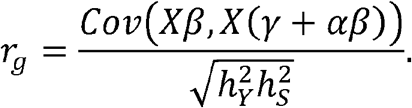

This allows an orthogonal decomposition of the SNP effects of*S*:

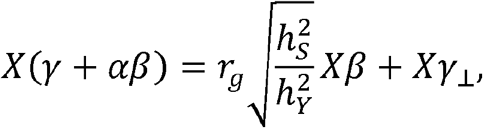

where*Xγ* _*⊥*_ is orthogonal to*Xβ;*.

In practice, the transformed phenotype*Y* ^′^ closely reflects a linear combination of target binary trait and*S*:

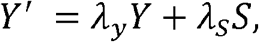

where*λ*_*y*_ and*λ*_*S*_ can be estimated by least squares after computing*Y* ^′^;. Substituting the orthogonal decomposition of*S* and rearranging:

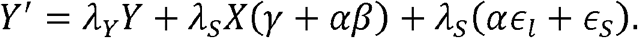

Using the decomposition above:

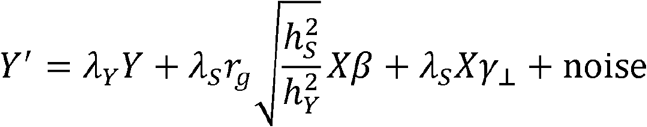

The critical term here is*λ*_*S*_*X γ* _⊥_, which represents genetic signal orthogonal to the liability of the target trait and thus constitutes a source of false positives.

Since

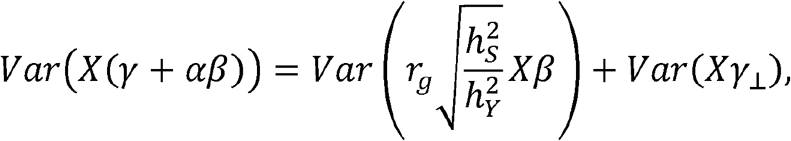

It follows that 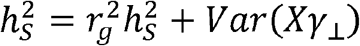 and therefore the variance of the false positive component is proportional to 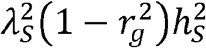, which we term the inflation criterion. To guard against false positives, StratGWAS requires by default that the inflation criterion does not exceed 0.01.

### Simulations

We simulated binary traits for 100,000 individuals using a liability threshold model, where individual liabilities were generated as

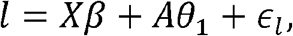

where*X* denotes the genotype,*β;* is the vector of SNP effect sizes,*A* is a vector containing the first principal component, θ_1_ is a scalar, and*ϵ*_*l*_ ∼*N* (0,*σ*^2^) is an independently distributed random noise term. The binary phenotype is defined by*Y* = 1_*l*>*τ*_, where threshold*τ* was selected to match prevalences of 0.2 and 0.5. We scaled*β;* so that the liability-scale heritability was either 0.2 or 0.5. The scalar θ_1_ was set such that the first principal component explained 5% of phenotypic variance on the liability scale.

For each simulated liability, we generated an associated stratification variable*S* with varying degrees of shared liability and genetic effects specific to*S*. Specifically, we modelled

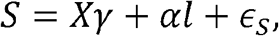

where*α* specified the degree of shared liability between*l* and*S, γ* specifies the SNP effect sizes of*S*, and*ϵ*_*S*_ is an independent noise term. We considered*α*= 0.2, 0.5, and 0.8 to capture low, moderate, and high degrees of shared liability, and heritabilities 0, 0.05, and 0.1 for the SNP effect sizes in*γ* specific to*S*, representing low, moderate, and high genetic correlation to target binary trait.

Causal SNP effects for*β;* were drawn from 5,000 randomly sampled SNPs on chromosomes 3–5, while we sampled 5,000 causal SNPs for*γ* from chromosomes 1–2. SNPs on chromosomes 6–22 served as null SNPs. The separation of causal SNPs for*β;* and*γ* allowed us to assess potential type 1 error inflation on chromosomes 1–2 arising from the use of*S* as stratification variable. All SNP effect sizes were sampled independently from a normal distribution and scaled to match the desired heritability.

