## Supplement for "Incorporating phenotype heterogeneity in disease GWAS improves power while maintaining specificity"

### Supplementary Notes


**1 Example StratGWAS commands** 4

**2 Description of StratGWAS** 6

2.1 Creating subgroups using input stratification variables . . . . . 6
2.2 Constructing the multi-phenotype matrix . . . . . 6
2.3 Subgroup-specific linear regression . . . . . 6
2.4 Computing LD scores . . . . . 7
2.5 Computing genetic covariance . . . . . 7
2.6 Computing the transformed phenotype . . . . . 8
2.7 Handling missing values of stratification variable . . . . . 9
2.8 Linear regression on transformed phenotype . . . . . 9

**3 Data** 10

3.1 Quality control of genotype data . . . . . 10
3.2 Selection of binary traits . . . . . 10
3.3 Age at onset and medication burden as stratification variables . . . . . 10
3.4 Stratification variables for major depressive disorder . . . . . 11

**4 Implementation of existing methods** 12

4.1 GenomicSEM . . . . . 12
4.2 ADuLT . . . . . 12

**5 Using a spline basis to model the transformed phenotype** 13

#### Supplementary Figures


1 Statistical power and type 1 error of StratGWAS versus case-control GWAS across different scenarios of stratification variables. . . . . 14
2 Type 1 error of StratGWAS versus case-control GWAS across different scenarios of stratification variables. . . . . 15
3 True underlying genetic liability versus StratGWAS transformed phenotype. . . . . 16
4 Transformed phenotype versus age-at-onset and medication burden. . . . . 17
5 Genetic covariance between subgroups of major depressive disorder. . . . . 18
6 StratGWAS weights of subgroups of major depressive disorder. . . . . 19


#### Supplementary Tables


1 Overview of 21 binary traits analyzed in the UK Biobank. . . . . 20
2 Overview of quintiles of age-at-onset. . . . . 21
3 Replication of SNPs in FinnGen. . . . . 22
4 Genetic correlations of case-control and StratGWAS-phenotypes between UK Biobank and FinnGen. . . 23
5 Overview of quintiles of medication uptake. . . . . 24
6 Summary and number of cases in strata of major depressive disorder. . . . . 25
7 Computational demands of StratGWAS. . . . . 26

### 1 Example StratGWAS commands

The StratGWAS R package constructs a transformed phenotype by stratifying disease cases into subgroups, and upweighting individuals carrying a higher genetic load. The following functions are sequentially run:

1. stratify — partition cases into strata;
2. compute_gencov — estimate genetic covariances between strata;
3. transform — generate the transformed phenotype;
4. linear — run a linear-regression GWAS on the transformed phenotype.

#### Input data format

All phenotype, covariate, and stratification files follow the standard PLINK 1 format: column 1 is the family ID (FID), column 2 is the individual ID (IID), and subsequent columns contain the relevant variables. Binary phenotypes are coded 0 (control) and 1 (case); missing values are coded NA. Genotype data should be provided in PLINK 1 (.bed/.bim/.fam) format.

#### Basic workflow

The code below illustrates the complete four-step workflow using a continuous stratification variable (e.g. age at diagnosis) together with an optional categorical variable (e.g. disease subtype) and a covariate file.

```
library(devtools)
install_github("JasperHof/StratGWAS")
library(StratGWAS)

# Load example data bundled with the package
data(pheno)       # Binary case-control phenotype
data(strat_cont)  # Continuous stratification variable
data(strat_cat)   # Categorical stratification variable
data(cov)         # Covariates (optional)

# Paths to PLINK genotype files (prefix without extension)
filename_bed <- system.file("extdata", "data.bed", package = "StratGWAS")
filename     <- gsub(".bed", "", filename_bed)

# Prefix for all output files
outfile <- tempfile("stratgwas_out")

# Step 1: stratify cases into subgroups
strata <- stratify(pheno, strat_cont = strat_cont, strat_cat = strat_cat, K = 5)
table(strata$info$groups)   # cases per stratum
head(strata$multi)          # multi-phenotype matrix
```

```
# Step 2: estimate genetic covariances between strata
gencov <- compute_gencov(strata, filename, nr_blocks = 1000, outfile)

gencov$gencov   # genetic covariance matrix
gencov$gencor   # genetic correlation matrix
gencov$hers     # per-stratum SNP heritabilities

# Step 3: build the transformed phenotype
trans <- transform(strata, gencov, outfile)

head(trans$transformed_pheno)   # transformed values
trans$weights                   # linear combination weights

# Step 4: GWAS on the transformed phenotype
linear(trans, filename, outfile, nr_blocks = 1000, cov = cov)
```

##### Output files

The table below summarizes the files written to disk using the supplied `outfile` prefix.

**Output files generated by `StratGWAS`.**

| **Function** | **File suffix** | **Contents** |
|---|---|---|
| `compute_gencov` | `.strata` | Multi-phenotype file for all strata |
| | `.pheno1`–`.phenoK` | Per-stratum GWAS summary statistics |
| | `.ldscores` | LD scores |
| | `.hers` | Per-stratum SNP heritabilities |
| | `.gencov` | Genetic covariance matrix |
| | `.gencor` | Genetic correlation matrix |
| `transform` | `.weights` | StratGWAS weights per subgroup |
| | `.transformed` | Transformed phenotype file |
| `linear` | `.assoc` | Final GWAS summary statistics |

### 2 Description of StratGWAS

#### 2.1 Creating subgroups using input stratification variables

Let $N$ denote the total number of individuals, $N_0$ the number of controls, and $N_1$ the number of cases. StratGWAS begins by stratifying the $N_1$ cases into subgroups according to one or more user-specified secondary phenotypes that are hypothesized to reflect disease genetic liability. We refer to these as "stratification variables", which may be continuous or categorical. Note that controls are not stratified and are shared across all subgroups.

For each continuous stratification variable $S$, StratGWAS begins by constructing a length-$N_1$ vector $S_1$ that contains the stratification variable values for all cases. The values in $S_1$ are ranked by numerical order and binned into $K$ approximately equal-sized groups of sizes $n_1, \ldots, n_K$ based on the empirical $0-$, $\frac{1}{K}-, \ldots, \frac{K-1}{K}-$, and $1-$quantiles of $S_1$. To ensure balanced sizes of subgroups in the presence of ties, a small normally distributed random jitter is added to $S_1$ before binning. Each case with non-missing $S_1$ is thus assigned to exactly one quantile-based subgroup per continuous stratification variable. Cases with missing stratification values $S_1$ are not assigned to specific subgroups but are later assigned to the mean value of cases with non-missing stratification variable $S_1$.

For each categorical stratification variable, the strata are defined directly by the observed categorical value. The number of groups is thus determined automatically from the unique values observed among cases, and each case is assigned to its corresponding category. Missing values are handled analogously to continuous variables. It is possible to run StratGWAS using multiple stratification variables. In this case, each input stratification variable results in its own set of strata, and strata from different variables are treated independently, rather than considering combinations of subgroups (which would grow exponentially with number of input variables).

#### 2.2 Constructing the multi-phenotype matrix

Suppose there are M input stratification variables, that yield subgroups of sizes $K_1, \ldots, K_M$. StratGWAS assembles all strata into a $N \times \sum_{j=1}^{M} K_j$-dimensional multi-phenotype matrix $Y$:

$$Y = (Y_{1,1}\, Y_{1,2}\, \ldots\, Y_{1,K_1}\, Y_{2,1}\, \ldots\, Y_{M,1}\, \ldots\, Y_{M,K_M})$$

Here, the length-$N$ vector $Y_{j,k}$ contains the stratified phenotype based on the $k$th subgroup of the $j$th stratification variable. For each individual $i$, these contain values:

$$(Y_{(j,k)})_i = \begin{cases} 1 & \text{case in subgroup } k \text{ of stratification variable } j \\ 0 & \text{control} \\ \text{missing} & \text{case in other subgroup than } k \text{ of stratification variable } j \end{cases}$$

#### 2.3 Subgroup-specific linear regression

Next, StratGWAS performs a linear regression for each phenotype in the multi-phenotype matrix using a simple linear regression. Note that StratGWAS does not regress out covariates from $Y_{j,k}$ by default (however, these are regressed out of the final transformed phenotype). All phenotypes are scaled to have mean 0 and variance 1 among non-missing observations, ensuring comparable effect estimates across phenotypes. To reduce memory footprint, genotypes are read in blocks of SNPs (default: 1 000 SNPs). For each SNP, StratGWAS computes the MAF and missingness rate, and standardizes phenotype and genotype within the set of individuals with non-missing data for both variables.

The regression coefficients are computed using a simple linear regression, with standard errors calculated based on the residual sum of squares. In addition, StratGWAS reports $\chi^2(1)$ statistics, along with p-values.

##### 2.4 Computing LD scores

StratGWAS continues to estimate LD scores for each SNP. By default, the LD score is computed as the sum of squared correlation ($r^2$) between a SNP and all other SNPs on the same chromosome located within 1 megabase (Mb) window. Genotypes are read in overlapping blocks for computational efficiency, with block size determined based on SNP density to ensure complete coverage of 1Mb windows.

##### 2.5 Computing genetic covariance

After performing stratum-specific linear regression and computing LD scores, StratGWAS estimates SNP heritabilities and genetic covariances between case strata using an implementation analogous to SumHer [1]. For each stratum, we estimate SNP heritability from summary statistics using the model

$$E(\chi_j^2) = 1 + N_j\, h^2_{\text{SNP}}\, q_j\, l_j + c$$

where $\chi_j^2$ is the observed chi-square statistic for SNP $j$, $N_j$ is the sample size, $l_j$ is the LD score, and $q_j = (\text{MAF}_j\,(1-\text{MAF}_j))^{1+\alpha}$ is the SNP prior with default $\alpha = -0.25$. The $q_j$ are normalized to sum to one across SNPs. The 'intercept' term $c$ can capture inflation due to confounding bias.

Parameters are estimated using iteratively reweighted least squares. At each iteration, StratGWAS performs a weighted regression on

$$\chi_j^2 - 1 = N_j\, h^2_{\text{SNP}}\, q_j\, l_j + cN_j + \epsilon_j$$

with weights derived from the inverse of the conditional variance, proportional to $(l_j\, \mu_j)^{-1}$, where $\mu_i$ denotes the current expectation. Expectations and weights are updated until convergence of a Gaussian pseudo-likelihood, and standard errors are reported based on a jack-knife estimator.

For pairs of strata A and B, we estimate genetic covariance using the product of Z-statistics. Let $Z_{j,A} = \frac{\beta_{j,A}}{\text{SE}(\beta_{j,A})}$ and $Z_{j,B} = \frac{\beta_{j,B}}{\text{SE}(\beta_{j,A})}$ denote Z-statistics for SNP $j$ in the two strata. We model

$$E(Z_{j,A}Z_{j,B}) = h^2_{AB}\, N_j^{\times}\, q_j\, l_j + c$$

where $N_j^{\times} = \sqrt{N_{j,A}N_{j,B}}$ is the geometric mean of sample sizes and $h_AB^2$ is the genetic covariance. The intercept term may absorb confounding bias or sample overlap. Estimation proceeds via iteratively reweighted least squares applied to

$$Z_{j,A}Z_{j,B} = h^2_{AB}\, N_j^{\times}\, q_j\, l_j + cN_j^{\times} + \epsilon_j$$

with weights proportional to $(l_j\, \mu_j)^{-1}$. Convergence is assessed using a Gaussian pseudo-likelihood [2], and standard errors are reported based on default standard errors from the weighted least squares regression model. Note that it is also possible to obtain jack-knife standard error estimates by specifying `jack = T` in the `compute_gencov` function.

Genetic correlations are finally computed and reported as

$$r_g = (h^2_{AB})/\sqrt{h^2_A h^2_B}.$$

#### 2.6 Computing the transformed phenotype

StratGWAS assembles the stratum-specific estimates of heritability and between-stratum genetic covariance estimates in a $\sum_{i=1}^{M} K_i \times \sum_{i=1}^{M} K_i$ genetic covariance matrix:

$$\Psi = \begin{pmatrix} h^2_{1,1} & \gamma_{(1,1),(1,2)} & \cdots & \gamma_{(1,1),(1,K_1)} & \gamma_{(1,1),(2,1)} & \cdots & \gamma_{(1,1),(M,K_M)} \\ \gamma_{(2,1),(1,1)} & h^2_{1,2} & \cdots & \vdots & & & \\ \vdots & \vdots & \ddots & \vdots & \vdots & & \\ \gamma_{(1,K_1),(1,1)} & \cdots & \cdots & h^2_{1,K_1} & & & \\ \gamma_{(2,1),(1,1)} & & & & h^2_{2,1} & & \\ \vdots & & & & & \ddots & \\ \gamma_{(M,K_M),(1,1)} & & & & & & h^2_{M,K_M} \end{pmatrix}$$

Next, for all stratification variables, the observed-scale heritabilities and genetic covariance estimates are transformed to liability-scale. The liablity scale heritability $h^2_l$ can be obtained from to observed-scale heritability $h^2_{\text{obs}}$ through the relationship [3]:

$$h^2_l = h^2_{\text{obs}} \cdot \frac{p(1-p)}{z^2} \tag{1}$$

where $z = \phi\big(\Phi^{-1}(1-p)\big)$ is the standard normal density evaluated at the liability threshold corresponding to prevalence $p$, with $\phi$ and $\Phi^{-1}$ denoting the standard normal PDF and inverse CDF, respectively. Note that this implicitly assumes that the observed prevalence matches the true prevalence, however this is not an issue for downstream analyses as this affects all heritability estimates equally.

We compute the liability scale heritablities $h^2_l$ for each stratum of each stratification variable based on their prevalence, and scale the corresponding rows and columns of $\Psi$ with $\sqrt{h^2_l/h^2_{\text{obs}}}$ such that all heritability and covariance estimates are represented on liability scale.

An eigendecomposition is then performed on $\Psi$, such that $\Psi = LDL^\top$, where $L = (L_{1,1}, L_{1,2}, \dots, L_{2,1}, \dots, L_{M,K_M})$ contains the eigenvectors and $D$ is the diagonal matrix. The first eigenvector $L_{1,1} = (l_{1,1}, l_{1,2}, \dots, l_{2,1}, \dots, l_{M,K_M})$ is a length-$\sum_{i=1}^{M} K_i$ vector, which is used as weights to compute the transformed phenotype. Prior to computing the transformed phenotype, $L_{1,1}$ is normalized and sign-adjusted (in case the mean value is negative) to ensure consistent interpretation. When specifying `jack = T`, StratGWAS obtains standard errors of $L_{1,1}$ by recomputing $L_{1,1}$ using the jack-knife estimates of heritability and genetic covariance estimates, which were computed in the previous step.

When analyzing categorical stratification variables only, the transformed phenotype is computed as a weighted sum across stratification variables:

$$Y = \sum_{j=1}^{M} \sum_{k=1}^{K_j} l_{j,k} Y'_{j,k}$$

where phenotype $Y'_{j,k}$ matches $Y_{j,k}$ except cases of subgroup $k$ of stratification variable $j$ are coded as 0, rather than missing.

When analyzing continuous stratification variables, StratGWAS approximates the relationship between stratification variable $S_j$ and stratum weights $l_{j,1}, \dots, l_{j,K_j}$ using a cubic spline. Suppose the $\frac{1}{K}$, ..., $\frac{K-1}{K}$−quantiles of $S_j$ are given by $t_{(1)}, ..., t_{(K-1)}$, such that subgroup $k$ contains all cases whose observed value $s$ of $S_j$ satisfies $t_{(k-1)} < s < t_{(k)}$, where $t_{(0)} = -\infty$ and $t_{(K)} = \infty$. Let $m_k$ denote the median of $S_j$ within each stratum $k$. StratGWAS fits a cubic spline $\hat{f}_j$ through the pairs $(m_1, l_{(j,1)}), ..., (m_K, l_{(j,K)})$, and evaluates it at each individual's observed value $s$ to obtain their smoothed transformed phenotype $\hat{f}_j(s)$.

In the case where at least one stratification variable is continuous, the individual-level transformed phenotype is assembled as a sum across stratification variables:

$$Y = \sum_{j=1}^{M} \hat{g}_j(s_{i,j}),$$

where $\hat{g}_j(s_{i,j})$ denotes the contribution of stratification variable $j$ to the transformed phenotype of individual $i$. For categorical stratification variables, $\hat{g}(s_{i,j}) = \sum_{k=1}^{K_j} l_{j,k} Y'_{j,k}$ as defined above. For continuous stratification variables, $\hat{g}_j(s_{i,j}) = \hat{f}_j(s_{i,j})$, where $\hat{f}_j$ is the fitted cubic spline for continuous stratification variable $j$.

#### 2.7 Handling missing values of stratification variable

If an individual is a case and missing for stratification variable $j$, StratGWAS imputes the transformed phenotype contribution from that variable using the mean transformed phenotype value across all non-missing cases for stratification variable $j$. Specifically, for continuous stratification variables, the mean of the spline-predicted weights across all non-missing cases is assigned. For categorical stratification variables, individuals missing for stratification variable $j$ are assigned the mean of $\sum_{k=1}^{K_j} l_{j,k} Y'_{j,k}$ among non-missing cases.

#### 2.8 Linear regression on transformed phenotype

The transformed phenotype $Y$ is first regressed on all provided covariates via ordinary least squares regression, with missing covariate values imputed to their column means. The residualized phenotype is then standardized to zero mean and unit variance called $\tilde{Y}$ and used for GWAS analysis. Note that this implementation is an approximate linear regression, as StratGWAS does not regress out covariates from the genotypes. If this is an issue, the user can provide the transformed phenotype in standard software such as PLINK [4], while if they prefer a mixed-model analysis and/or use of saddlepoint approximation, they can use LDAK-KVIK [5].

GWAS analysis is performed by reading in genotype data in SNP blocks (default block size: 1 000). Within each block, genotypes are mean-centered and scaled to unit variance, with missing genotype calls set to zero after centering. For each SNP $j$, the effect size estimate and its standard error are obtained from simple linear regression of the standardized phenotype $\tilde{Y}$ on the standardized SNP vector $g_j$:

$$\hat{\beta}_j = \frac{g_j^\top \tilde{Y}}{g_j^\top g_j}$$

$$\widehat{\mathrm{SE}}(\beta_j) = \sqrt{\frac{\mathrm{RSS}_j/(n_j - 2)}{g_j^\top g_j}}$$

where $\mathrm{RSS}_j = \tilde{Y}^\top \tilde{Y} - \hat{\beta}_j \cdot g_j^\top \tilde{Y}$ is the residual sum of squares and $n_j$ is the number of individuals with non-missing genotype and phenotype for SNP $j$. The chi-squared test statistic is computed as $\chi_j^2 = \left(\frac{\hat{\beta}_j}{\widehat{\mathrm{SE}}(\beta_j)}\right)^2$, and evaluated on one degree of freedom to obtain association p-values.

### 3 Data

#### 3.1 Quality control of genotype data

In this study, we used a subset of data from the UK Biobank containing 367 981 individuals, which were described previously in the Supplemental Notes of the LDAK-KVIK publication [5]. Briefly, after excluding individuals who had withdrawn consent or were missing phenotype information, we restricted to the 408 868 individuals self-identifying as white British with confirmed similar genetic ancestry based on principal component analysis (UK Biobank data field 22006). From these, we randomly selected 367,981 individuals (90%) as our primary analysis dataset. We restricted to autosomal, biallelic, directly genotyped SNPs with MAF $> 0.001$ and genotype call rate $> 90\%$, yielding 690 264 SNPs; no filtering on Hardy-Weinberg equilibrium or genotype rate was applied.

For our simulations, we used a subset of the 408 868 White British individuals described above. We additionally filtered on individuals with age, sex, and townsend deprivation recorded for each individual, and applied a relatedness filter such that no two individuals were more closely related than third-degree relatives. This yielded 359 444 individuals, from whom we selected 100 000 for use in simulation studies. For our simulations dataset, we restricted to autosomal, biallelic, directly genotyped SNPs with MAF $> 0.01$ and genotype call rate $> 90\%$, yielding 641 204 SNPs.

#### 3.2 Selection of binary traits

For our application on UK Biobank traits, we restricted to diseases recorded using 3-digit ICD-10 codes (category 1712), as these provided information on date of first occurrence and could thus be used to derive age at onset. This yielded 1 140 diseases, of which 74 had a prevalence exceeding 5% in our analysis dataset. We next performed a case-control GWAS for each of these 74 diseases, including the first 10 principal components, age, age$^2$, sex, and age$\times$sex as covariates, and estimated observed-scale SNP heritability from the summary statistics using SumHer [1]. In total, 21 diseases with observed scale heritability exceeding 0.05 were carried forward for analysis.

#### 3.3 Age at onset and medication burden as stratification variables

Age at onset was derived from the date of first occurrence of each ICD-10 diagnosis (UK Biobank category 1712), computed as the difference in years between date of diagnosis and date of birth. As only birth year and birth month are available in the UK Biobank, day of birth was imputed as the 15th day of the month.

Medication burden was constructed using primary care prescription data available for 180,026 UK Biobank participants [6]. For each disease, we identified medications belonging to the British National Formulary (BNF) chapters corresponding to the ICD-10 chapter of the disease, using the following mapping of BNF chapter:

- E: Infections
- F: Central nervous system
- I: Cardiovascular system
- J: Respiratory system
- K: Gastro-intestinal system

- M: Musco skeletal and joint diseases

Medication burden was defined as the number of unique medications prescribed within the matched BNF category, divided by the individual's age at the time of data collection to account for the accumulation of prescriptions over time.

For the comparison between StratGWAS using medication burden and other methods, we restricted to the analyses of the subset of 136 304 individuals among the 367 901 individuals for whom primary care prescription data were available.

#### 3.4 Stratification variables for major depressive disorder

We considered five categories of stratification variables for major depressive disorder (MDD), which we extracted from the UK Biobank using the DNAnexus Research Analysis Platform.

The first category captured the source of depression diagnosis, derived by combining two self-report fields: whether the participant had seen a doctor for nerves, anxiety, tension or depression (field 2090) and whether they had seen a psychiatrist for such difficulties (2100). These were combined into a three-category variable: doctor only, psychiatrist only, or both.

The second category captures electronic health record-based diagnosis, using ICD-10 codes F32 (depressive episode; field 130894) and F33 (recurrent depressive disorder; field 130896), coded as binary indicators of whether a diagnosis was recorded.

The third category captured whether the cases had ever been offered or sought treatment for depression (field 21063), which was coded as binary indicator.

The fourth category captured psychiatric comorbidities, specifically the presence of a recorded diagnosis of bipolar disorder (field 130892) and schizophrenia (field 130874), each coded as binary indicator.

The fifth category captured self-reported depressive symptoms over the past two weeks, comprising four items previously associated with MDD subtypes [7]: depressed mood (field 2050), disinterest (field 2060), restlessness (field 2070), and tiredness (field 2080). Responses were coded on a four-point ordinal scale: not at all, several days, more than half the days, and nearly every day. Negative values indicating as missing or prefer-not-to-answer responses were set to missing prior to analysis.

All stratification variables were assembled at the individual level and merged with the primary MDD phenotype file for input into StratGWAS.

### 4 Implementation of existing methods

#### 4.1 GenomicSEM

We applied GenomicSEM [8] to perform multivariate GWAS analysis using subgroup-specific summary statistics. These subgroup-specific summary statistics were generated in the second step of StratGWAS, where we stratified individuals based on a continuous variable and performed separate GWAS within each stratum. In the UK Biobank application, the subgroups were defined based on quintiles of age at onset or medication uptake.

The GenomicSEM analysis proceeded in four steps. First, we munged the subgroup-specific summary statistics files using the `munge` function, restricting to HapMap3 SNPs and specifying the sample size for each subgroup. Second, we performed multivariable LD score regression (LDSC) using the `ldsc` function to estimate the genetic covariance structure between subgroups. For LD scores, we used those provided by the original developers of LDSC, based on a European population and excluding MHC region. We specified sample prevalence and population prevalence based on the observed proportions of cases in each subgroup within our study sample. Third, we prepared the GWAS summary statistics using the `sumstats` function, applying MAF filtering (MAF $> 0.01$) and excluding indels. Finally, we performed the multivariate GWAS using the `userGWAS` function, specifying a common factor model where a single latent factor F1 loaded onto all five subgroup phenotypes, and this factor was regressed on each SNP. We used the options `GC='none'` to disable genomic control, `smooth_check=TRUE` to check for non-positive definite matrices, and `fix_measurement=TRUE` to fix factor loadings.

#### 4.2 ADuLT

Prior to running ADuLT [9], we computed familial relationship among UK Biobank participants using KING software [10], resulting in 4 323 parent-offspring pairs, 15 648 full sibling pairs, and 124 pairs of monozygotic twins. We next formatted our individual age-at-onset data into a format suitable for ADuLT and computed cumulative incidence curves after stratifying individuals based on 5-years periods of birth (e.g., 1940-1944, 1945-1949, etc.) and sex. The familial relationships were specified using the relationships obtained from KING software, and finally used the `estimate_liability` function to compute individual liabilities.

We next analyzed the ADuLT-derived phenotype in a linear regression in LDAK software, using the same covariates used for StratGWAS transformed phenotype.

### 5 Using a spline basis to model the transformed phenotype

In an earlier implementation of StratGWAS, we explored whether secondary phenotypes could be incorporated more directly by constructing a genetically informed composite trait. This approach enables a more direct modelling of genetic liability as a function of stratification variable but can introduce false positives in some scenarios. We explain this approach below.

Let $N$ denote the total number of individuals, $Y$ a binary trait of interest, and $Z$ a stratification variable (e.g., age at diagnosis, medication burden, etc.). After centering and scaling $Y$ and $Z$, it is possible to construct a multivariate phenotype

$$A = (Y, B(Z)),$$

where $B(Z)$ denotes a basis for modelling the genetic liability as function of $Z$. We previously implemented a 5-dimensional spline basis $B(Z) = (Z, Z^2, Z^3, (Z - k_1(Z))^3, (Z - k_2(Z))^3)$, where $k_1(Z)$ and $k_2(Z)$ denote the $\frac{1}{3}$- and $\frac{2}{3}$-quantiles of Z.

We can model $A$ under a multivariate linear mixed model with genetic and environmental covariance components:

$$A \sim N(X\beta, \Psi \otimes K + \Phi \otimes I_N),$$

where $X$ the genotype matrix, $\beta$ denotes SNP effect sizes across phenotypic components, $K$ is the genomic relatedness matrix (GRM), $I_N$ is the $N \times N$ identity matrix, and $\Psi, \Phi$ represent the genetic and environmental covariance matrices among the components of $A$, respectively.

It is then possible to estimate $\Psi$ and $\Phi$ using randomized Haseman-Elston regression. Given these estimates, a linear transformation $L$ can be computed that simultaneously diagonalizes the genetic and environmental covariance [11], satisfying

$$L\Psi L^\top = D, \qquad L\Phi L^\top = I,$$

where $D$ is diagonal and $I$ denotes the identity matrix.

We can then transform the phenotype matrix as $A' = AL$, which follows

$$A' \sim N(X\beta\, L, D \otimes K + I_N),$$

By construction, the eigenvalues in $D$ are ordered decreasingly, and the first eigenvector defined the linear combination of $(Y, B(Z))$ that maximizes the ratio of genetic to environmental variance, i.e., the heritability. Consequently, the first transformed phenotype component, $A'_1 = Al_1$, where $l_1$ is the leading column of $L$, represents the direction of maximal genetic signal across strata. StratGWAS uses $A'_1$ as the outcome phenotype in GWAS analysis.

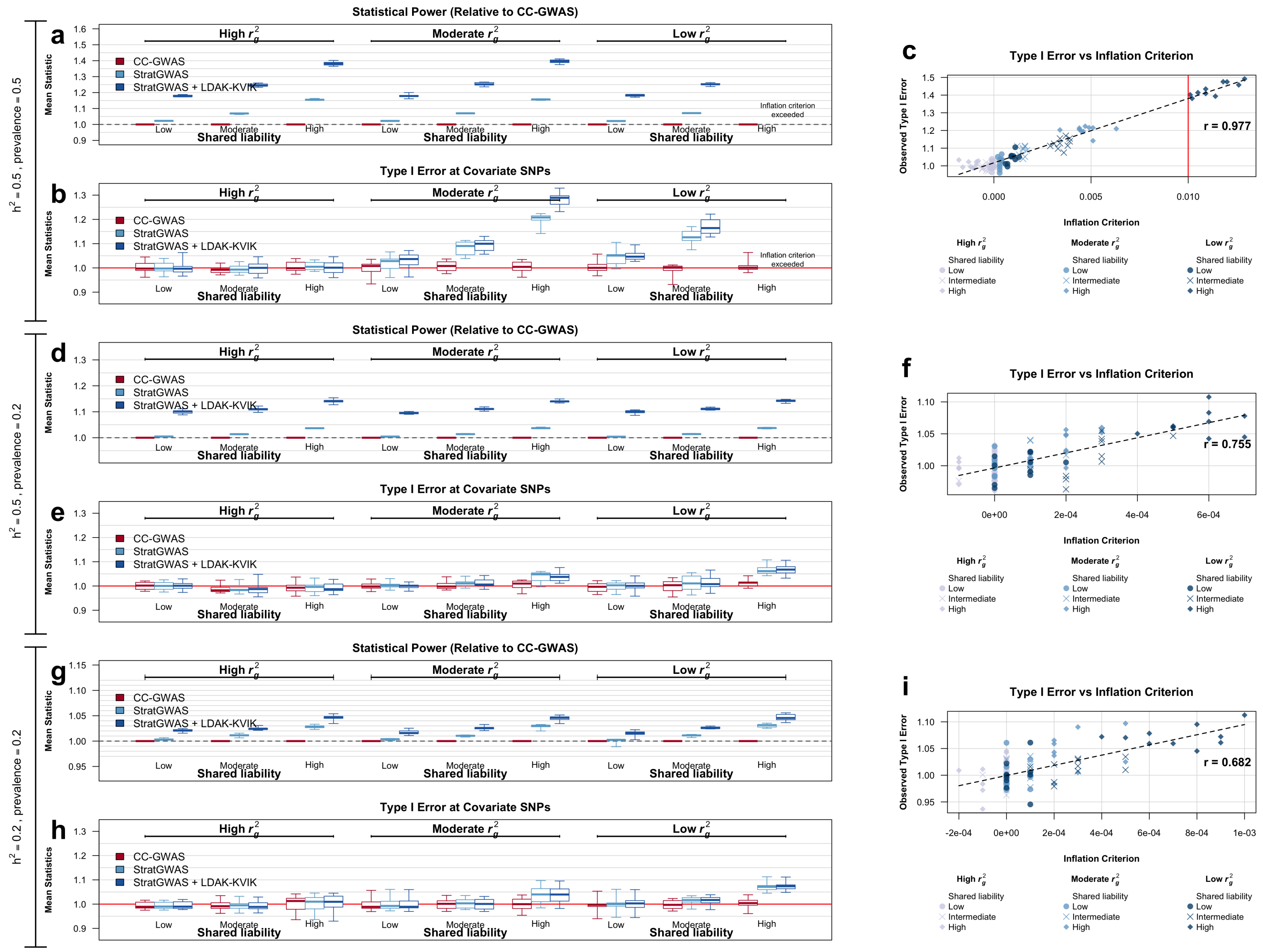


**Supplementary Figure 1: Statistical power and type 1 error of StratGWAS versus case-control GWAS across different scenarios of stratification variables.**

We generated binary traits with varying prevalence and liability heritability for 100 000 individuals. Ten phenotypes were simulated for each scenario, which and an additional nine associated stratification variable were generated for each binary phenotype, varying in shared liability and genetic correlation with target trait. Panels report the mean $\chi^2(1)$ test statistic of causal SNPs relative to case-control GWAS (**a, d, g**), and mean $\chi^2(1)$ test statistic of SNPs that are causal to the stratification variable, but not the binary trait (**b, e, h**). Panels on the right (**c, f, i**) compare the estimated inflation criterion against observed type 1 errors for all simulated combinations, with the fitted correlation shown on right. The red vertical line at $x = 0.01$ represents the default inflation criterion threshold; results are not shown for scenarios exceeding this threshold, as StratGWAS is not recommended in these settings.

Across all simulation scenarios considered, power improved with increased shared liability, while type 1 errors increased whenever genetic correlation was low and shared liability was high. Largest power gains of StratGWAS were observed when prevalence was highest, while the benefit of integrating the StratGWAS-transformed phenotype with LDAK-KVIK was highest for phenotypes with high heritability. The inflation criterion correlated highly with observed type 1 error rates, identifying one scenario where confounding by stratification variable introduced false positives.

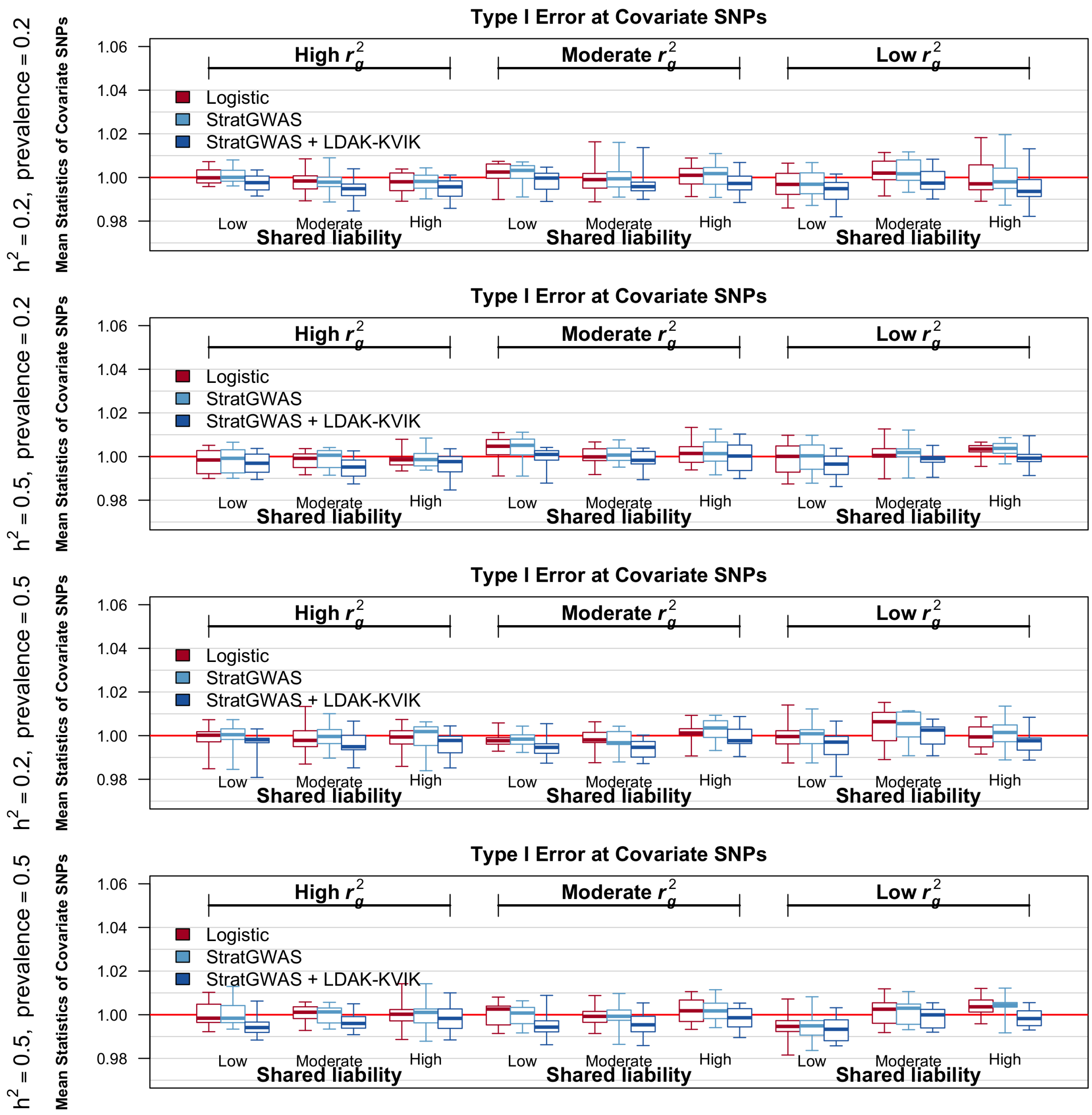


**Supplementary Figure 2: Type 1 error of StratGWAS versus case-control GWAS across different scenarios of stratification variables.**

We generated binary traits with varying prevalence and liability heritability for 100 000 individuals. Ten phenotypes were simulated for each scenario, which and an additional nine associated stratification variable were generated for each binary phenotype, varying in shared liability and genetic correlation with target trait. Panels report the mean $\chi^2(1)$ test statistic of null SNPs that were noncausal to both the target trait and stratification variable.

Across all scenarios considered, StratGWAS and case-control GWAS controlled type 1 errors at null SNPs that were noncausal to both target trait and stratification variable.

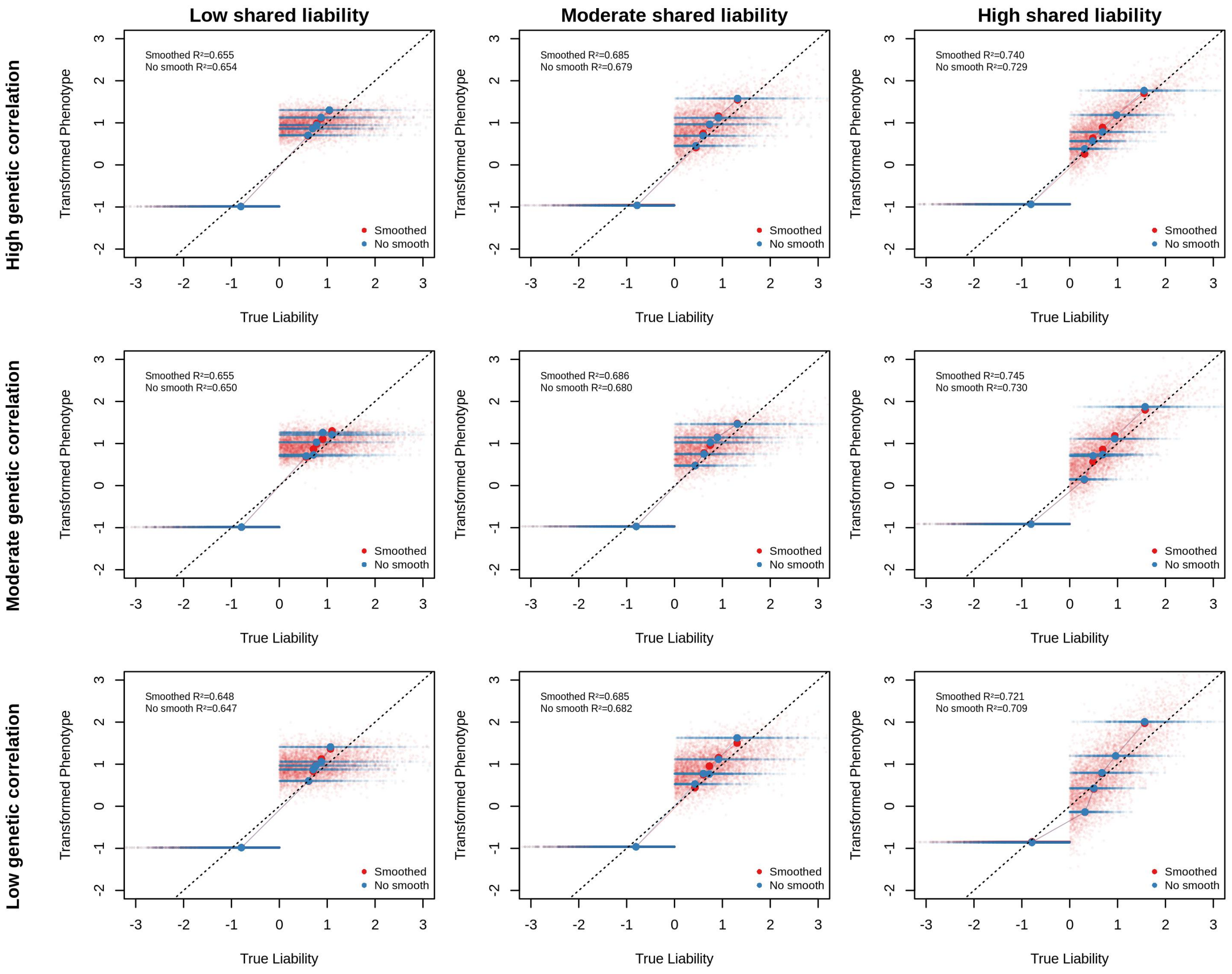


**Supplementary Figure 3: True underlying genetic liability versus StratGWAS transformed phenotype.** We simulate binary phenotypes of heritability 0.2 and prevalence 0.5 for 100 000 individuals, and generated nine associated stratification variables, that shared varying degrees of liability and genetic correlation to target trait. Each combination of phenotype and stratification variable was analyzed using StratGWAS, both with and without smoothing to model the relationship between stratification variable and transformed phenotype. Each plot represents a different simulation scenario, with small points representing individual-level comparisons of true underlying liability versus StratGWAS transformed phenotype. Dots compare the average true liability with average transformed phenotype across each of five subgroups defined by quintiles of the stratification variable.

We find that the StratGWAS transformed phenotypes accurately reflect underlying liability, with increasing accuracy when there is larger shared liability between target trait and stratification variable. Moreover, transformed phenotypes are most accurate when in case of high genetic correlation between target trait and stratification variable, and is further improved by modelling the relationship between stratification variable and transformed phenotype using smoothing.

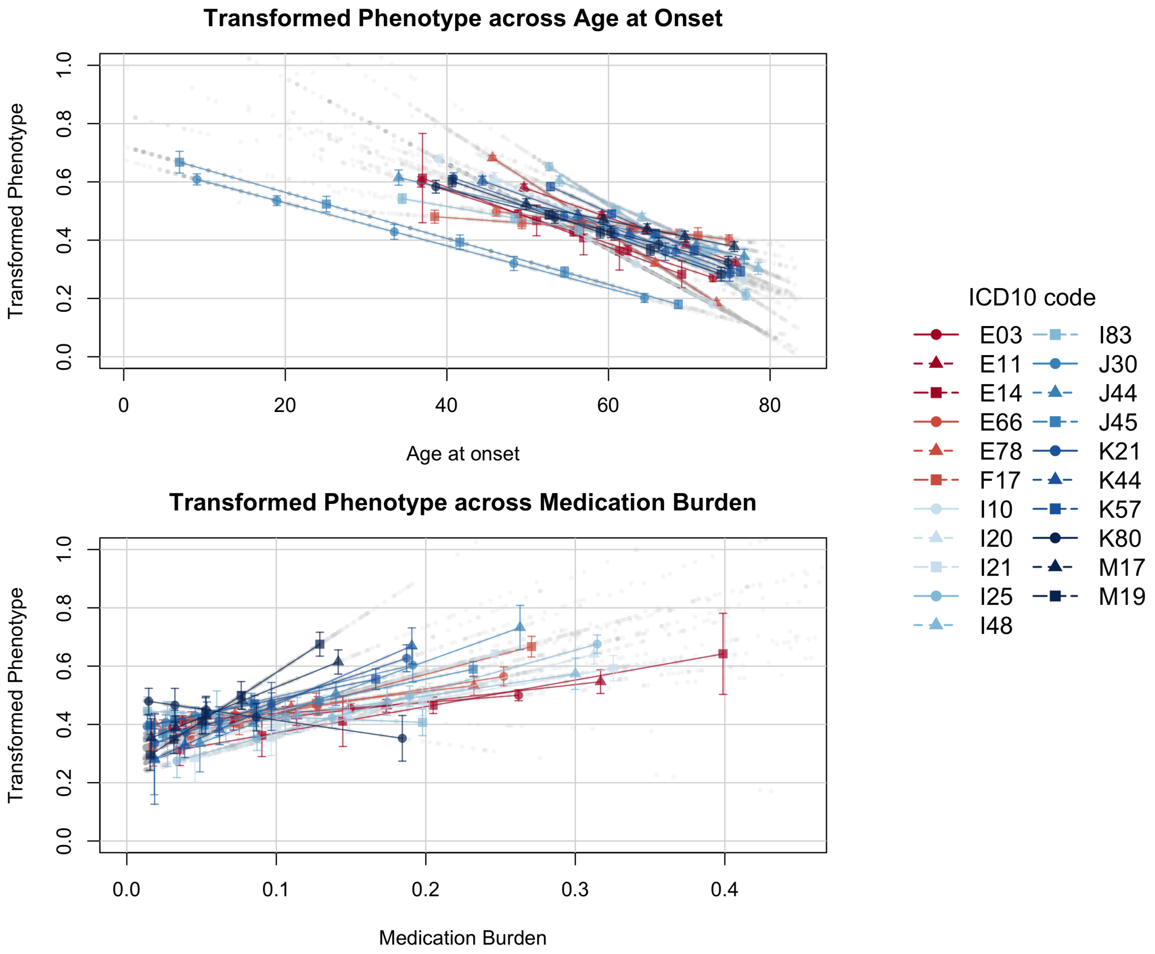


**Supplementary Figure 4: Transformed phenotype versus age-at-onset and medication burden.** We computed transformed phenotypes for 21 traits recored in the UK Biobank, using age-at-onset and medication burden as continuous stratification variables. Plots display the value of the StratGWAS-transformed phenotype versus age-at-onset and medication burden. Dots represent the mean value of age-at-onset versus the transformed phenotype across quintiles of the stratification variable, with standard errors of the StratGWAS weights.

Across all traits considered, StratGWAS tended to assign higher weight to individuals in subgroups of early onset, or those characterized by higher medication uptake.

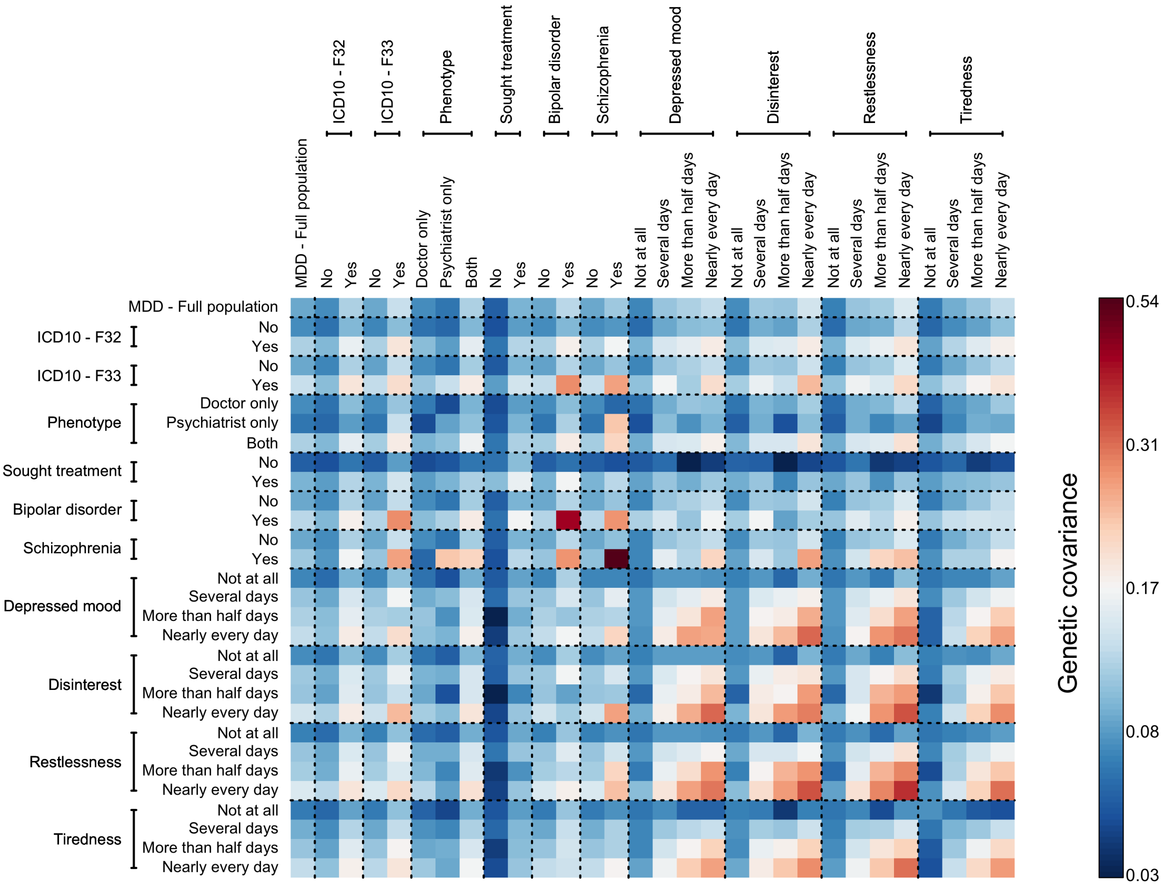


**Supplementary Figure 5: Genetic covariance between subgroups of major depressive disorder.** After stratifying MDD cases in subgroups in the first step of StratGWAS, we computed the genetic covariance among disease subgroups. The colored boxes represent the strength of the genetic covariance between subgroups, here presented on liability scale, ranging from dark blue (low genetic covariance) to dark red (high genetic covaraince). Values on the diagonal represent the liability scale heritablity of subgroups.

For all stratification variables considered, subgroups showing multiple modes of diagnosis, comorbidity, or more depressive symptoms, were associated with higher heritability and genetic covariance with other subgroups.

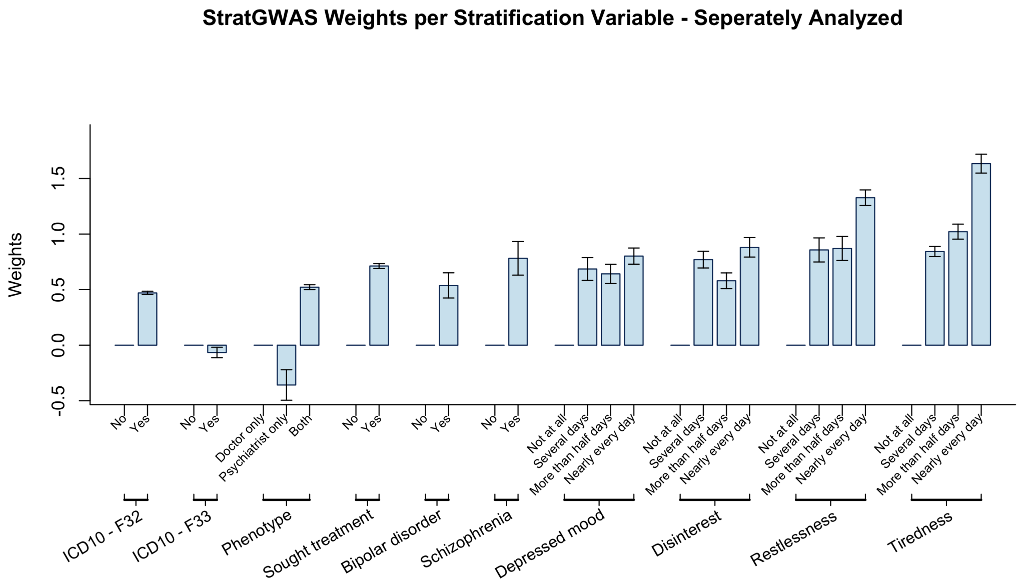


**Supplementary Figure 6: StratGWAS weights of subgroups of major depressive disorder.** We analyzed major depressive disorder in the UK Biobank in StratGWAS using different stratification variables, based on different types of diagnoses, comorbidities, and depression-related symptoms. While the main paper analyzes all categorical stratification variables jointly in StratGWAS, we here present the stratification weights obtained from analyzing each stratification variable individually. Weights represent the additional assigned phenotypic value to individuals in that category with respect to reference group, where the phenotype of controls are coded as 0, the reference subgroup as 1, and individuals in other subgroups as 1 + Weight.

Similar as to the joint analysis, individuals with multiple modes of diagnosis, comorbidity or depression-related symptoms were assigned higher weights in the StratGWAS-transformed phenotype compared to those without. Analyzing each stratification variable separately resulted in much higher weights compared to analyzing them jointly, which is likely explained by correlation among disease strata, resulting in lower marginal effects in the joint analysis.

| Trait | ICD-10 code | N (prevalence) | $h^2_{\text{liab}}$ |
|---|---|---|---|
| Hypothyroidism, unspecified | E03 | 40429 (8.1) | 0.34 |
| Type 2 diabetes mellitus | E11 | 47069 (9.4) | 0.24 |
| Unspecified diabetes mellitus | E14 | 26312 (5.2) | 0.23 |
| Obesity | E66 | 53064 (10.6) | 0.14 |
| Disorders of lipoprotein metabolism and other lipidemias | E78 | 128634 (25.6) | 0.15 |
| Mental and behavioural disorders due to use of tobacco | F17 | 45825 (9.1) | 0.11 |
| Essential (primary) hypertension | I10 | 203377 (40.5) | 0.19 |
| Angina pectoris | I20 | 38647 (7.7) | 0.13 |
| Acute myocardial infarction | I21 | 25198 (5.0) | 0.15 |
| Chronic ischaemic heart disease | I25 | 56044 (11.2) | 0.13 |
| Atrial fibrillation and flutter | I48 | 42863 (8.5) | 0.13 |
| Varicose veins of lower extremities | I83 | 25600 (5.1) | 0.18 |
| Vasomotor and allergic rhinitis | J30 | 52006 (10.4) | 0.13 |
| Other chronic obstructive pulmonary disease | J44 | 28185 (5.6) | 0.13 |
| Asthma | J45 | 74385 (14.8) | 0.19 |
| Gastro-oesophageal reflux disease | K21 | 82285 (16.4) | 0.08 |
| Diaphragmatic hernia | K44 | 61861 (12.3) | 0.10 |
| Diverticular disease of intestine | K57 | 74399 (14.8) | 0.13 |
| Cholelithiasis | K80 | 35894 (7.1) | 0.12 |
| Gonarthrosis [arthrosis of knee] | M17 | 48361 (9.6) | 0.12 |
| Other and unspecified osteoarthritis | M19 | 92720 (18.5) | 0.08 |

**Supplementary Table 1: Overview of 21 binary traits analyzed in the UK Biobank.** We selected traits based on 3-digit ICD-10 codes in the UK Biobank that had prevalence >5% were estimated to have observed-scale heritability estimate >5% using SumHer [1].

| | | Quantile of age-at-onset | | | | |
|---|---|---|---|---|---|---|
| ICD-10 code | N cases (prevalence) | 20% | 40% | 60% | 80% | Inflation criterion |
| E03 | 40429 (8.1) | 44.71 | 52.37 | 58.54 | 66.24 | 0.0114 |
| E11 | 47069 (9.4) | 56.15 | 62.15 | 67.23 | 72.33 | 0.0031 |
| E14 | 26312 (5.2) | 47.38 | 54.46 | 59.38 | 64.13 | 0.0459 |
| E66 | 53064 (10.6) | 52.75 | 60.02 | 66.01 | 71.80 | 0.0002 |
| E78 | 128634 (25.6) | 52.71 | 58.62 | 63.21 | 68.64 | 0.0050 |
| F17 | 45825 (9.1) | 46.03 | 52.95 | 59.41 | 66.66 | 0.0002 |
| I10 | 203377 (40.5) | 47.47 | 54.55 | 60.46 | 67.13 | 0.0055 |
| I20 | 38647 (7.7) | 52.45 | 58.46 | 63.70 | 70.00 | 0.0017 |
| I21 | 25198 (5) | 51.54 | 58.54 | 64.66 | 71.51 | 0.0022 |
| I25 | 56044 (11.2) | 57.99 | 63.57 | 68.33 | 73.33 | 0.0034 |
| I48 | 42863 (8.5) | 60.68 | 66.93 | 71.38 | 75.60 | -0.0006 |
| I83 | 25600 (5.1) | 44.26 | 53.09 | 59.58 | 66.91 | 0.0004 |
| J30 | 52006 (10.4) | 13.96 | 25.46 | 40.47 | 55.46 | 0.0047 |
| J44 | 28185 (5.6) | 54.99 | 62.71 | 68.38 | 73.51 | 0.0017 |
| J45 | 74385 (14.8) | 14.46 | 35.46 | 48.47 | 60.38 | 0.0255 |
| K21 | 82285 (16.4) | 50.31 | 57.65 | 63.63 | 70.26 | 0.0011 |
| K44 | 61861 (12.3) | 52.46 | 59.50 | 65.51 | 71.42 | 0.0020 |
| K57 | 74399 (14.8) | 57.05 | 63.38 | 68.28 | 73.00 | 0.0000 |
| K80 | 35894 (7.1) | 49.13 | 56.71 | 62.90 | 69.77 | 0.0015 |
| M17 | 48361 (9.6) | 56.20 | 62.17 | 66.97 | 71.98 | 0.0003 |
| M19 | 92720 (18.5) | 49.04 | 56.11 | 61.96 | 69.17 | 0.0020 |

**Supplementary Table 2: Overview of quintiles of age-at-onset.**

For our application of StratGWAS to age-at-onset, we stratified cases into quintiles, i.e., using the $20\%-, 40\%-, 60\%-$, and 80%-quantiles of age-at-onset as cut-off points for subgroups. Columns present these cut-off points in age at diagnosis in years. While the quantiles of most diseases were observed between age 50 and 70, some of the diseases of the respiratory system were characterized by early onset.

| | | | | Replication threshold | | |
|---|---|---|---|---|---|---|
| Analysis | Method | Total sign. | No. indep. | 0.05 | 0.05/(No. indep.) | $5 \cdot 10^{-8}$ |
| Age-at-onset | CC-GWAS | 12485 | 1361 | 1140 (84%) | 918 (67%) | 592 (43%) |
| | StratGWAS | 14825 | 1588 | 1292 (81%) | 985 (62%) | 615 (39%) |
| | StratGWAS + LDAK-KVIK | 14644 | 1636 | 1334 (82%) | 1015 (62%) | 635 (39%) |
| | GenomicSEM | 11673 | 1351 | 1135 (84%) | 913 (68%) | 598 (44%) |
| | ADuLT | 14259 | 1552 | 1260 (81%) | 972 (63%) | 613 (39%) |
| Medication burden | CC-GWAS | 3378 | 299 | 272 (91%) | 258 (86%) | 208 (70%) |
| | StratGWAS | 3717 | 310 | 276 (89%) | 257 (83%) | 208 (67%) |
| | StratGWAS + LDAK-KVIK | 3862 | 342 | 305 (89%) | 281 (82%) | 227 (66%) |
| | GenomicSEM | 3135 | 282 | 258 (91%) | 242 (86%) | 205 (73%) |

**Supplementary Table 3: Replication of SNPs in FinnGen.**

We replicated the results from analyzing 21 binary traits in the UK Biobank using summary statistics from FinnGen [12], which were available for all traits except for ICD-10 codes F17 and K44. Columns present the number of genome-wide signficant SNPs and independent significant SNPs for all methods considered, followed by their replication rates in FinnGen. Note that the application of methods within the 'Age-at-onset' category were based on 367 981 individuals, while the we compared methods in the "Medication burden" category based on the subset of 136 304 individuals with medication data available. Numbers of replicated SNPs were evaluated based on number of independent significant SNPs exceeding P value thresholds $0.05, 0.05/n_{\text{sign}}$, and $5 \cdot 10^{-8}$, where $n_{\text{sign}}$ indicates the number of independent significant SNPs for the respective method, evaluated for each phenotype.

For both applications, integrating the StratGWAS-transformed phenotype in LDAK-KVIK yielded the highest number of independent genome-wide significant SNPs, and resulted in highest number of replicated loci across replication thresholds. ADuLT and StratGWAS yielded similar numbers of significant and replicated SNPs, while those from GenomicSEM were slightly behind case-control GWAS. Overall, the high replication rates from StratGWAS suggest that the transformed phenotypes maintain disease specificity while improving statistical power.

| | Age-at-onset application | | Medication burden application | |
|---|---|---|---|---|
| ICD-10 code | CC-GWAS | StratGWAS | CC-GWAS | StratGWAS |
| E03 | 0.80 (0.77, 0.84) | 0.80 (0.77, 0.84) | 0.79 (0.74, 0.85) | 0.79 (0.73, 0.85) |
| E11 | 0.85 (0.82, 0.88) | 0.85 (0.82, 0.88) | 0.90 (0.85, 0.95) | 0.90 (0.85, 0.95) |
| E14 | 0.86 (0.82, 0.89) | 0.86 (0.82, 0.89) | 0.86 (0.79, 0.93) | 0.84 (0.76, 0.92) |
| E66 | 0.81 (0.77, 0.85) | 0.81 (0.77, 0.85) | 0.82 (0.76, 0.88) | 0.83 (0.77, 0.89) |
| E78 | 0.85 (0.74, 0.96) | 0.84 (0.73, 0.95) | 0.82 (0.69, 0.95) | 0.82 (0.69, 0.95) |
| I10 | 0.85 (0.83, 0.87) | 0.85 (0.83, 0.87) | 0.87 (0.83, 0.90) | 0.86 (0.83, 0.89) |
| I20 | 0.86 (0.82, 0.91) | 0.86 (0.81, 0.90) | 0.86 (0.78, 0.94) | 0.84 (0.76, 0.91) |
| I21 | 0.88 (0.84, 0.93) | 0.88 (0.83, 0.93) | 0.87 (0.79, 0.96) | 0.86 (0.77, 0.95) |
| I25 | 0.88 (0.84, 0.91) | 0.87 (0.84, 0.91) | 0.92 (0.85, 0.99) | 0.90 (0.84, 0.97) |
| I48 | 0.85 (0.80, 0.90) | 0.84 (0.80, 0.89) | 0.82 (0.74, 0.90) | 0.85 (0.77, 0.92) |
| I83 | 0.87 (0.83, 0.92) | 0.87 (0.83, 0.92) | 0.87 (0.80, 0.94) | 0.83 (0.75, 0.91) |
| J30 | 0.74 (0.65, 0.82) | 0.73 (0.65, 0.81) | 0.73 (0.63, 0.84) | 0.76 (0.67, 0.86) |
| J44 | 0.63 (0.58, 0.68) | 0.63 (0.59, 0.68) | 0.66 (0.58, 0.75) | 0.70 (0.62, 0.77) |
| J45 | 0.80 (0.77, 0.84) | 0.77 (0.74, 0.81) | 0.82 (0.76, 0.87) | 0.83 (0.78, 0.89) |
| K21 | 0.80 (0.73, 0.87) | 0.80 (0.73, 0.87) | 0.81 (0.70, 0.91) | 0.82 (0.72, 0.92) |
| K57 | 0.90 (0.86, 0.94) | 0.89 (0.86, 0.93) | 0.86 (0.79, 0.92) | 0.86 (0.80, 0.93) |
| K80 | 0.89 (0.85, 0.94) | 0.90 (0.85, 0.94) | 0.92 (0.83, 1.01) | 0.91 (0.82, 0.99) |
| M17 | 0.81 (0.77, 0.86) | 0.81 (0.77, 0.86) | 0.83 (0.76, 0.90) | 0.83 (0.76, 0.90) |
| M19 | 0.78 (0.72, 0.83) | 0.77 (0.71, 0.83) | 0.75 (0.66, 0.83) | 0.75 (0.67, 0.83) |
| **Mean** | **0.83** | **0.82** | **0.83** | **0.83** |

**Supplementary Table 4: Genetic correlations of case-control and StratGWAS-phenotypes between UK Biobank and FinnGen.**

We computed the genetic correlation of 19 traits based on case-control GWAS and StratGWAS with summary statistics available from FinnGen [12]. Genetic correlations are here reported with 95% confidence interval, and were computed using SumHer [1], using a subset of 1 000 individuals from the 367 981 UK Biobank participants as reference panel.

Across all traits, the genetic correlations between case-control GWAS and FinnGen were highly similar to that of StratGWAS and FinnGen, suggesting similar degrees of genetic overlap between the two phenotypes. This also suggests that the StratGWAS-transformed phenotypes maintained disease specificity.

| | | Quantile of medication burden | | | | |
|---|---|---|---|---|---|---|
| ICD-10 code | N cases (prevalence) | 20% | 40% | 60% | 80% | Inflation criterion |
| E03 | 40429 (8.1) | 0.05 | 0.09 | 0.13 | 0.18 | 0.0007 |
| E11 | 47069 (9.4) | 0.04 | 0.08 | 0.13 | 0.22 | 0.0031 |
| E14 | 26312 (5.2) | 0.06 | 0.11 | 0.17 | 0.26 | 0.0218 |
| E66 | 53064 (10.6) | 0.03 | 0.06 | 0.10 | 0.16 | 0.0014 |
| E78 | 128634 (25.6) | 0.03 | 0.05 | 0.08 | 0.14 | 0.0014 |
| F17 | 45825 (9.1) | 0.01 | 0.04 | 0.08 | 0.15 | 0.0113 |
| I10 | 203377 (40.5) | 0.02 | 0.06 | 0.10 | 0.15 | 0.0018 |
| I20 | 38647 (7.7) | 0.06 | 0.11 | 0.16 | 0.23 | 0.0027 |
| I21 | 25198 (5.0) | 0.04 | 0.11 | 0.16 | 0.23 | 0.0021 |
| I25 | 56044 (11.2) | 0.04 | 0.10 | 0.15 | 0.22 | 0.0024 |
| I48 | 42863 (8.5) | 0.03 | 0.07 | 0.13 | 0.20 | 0.0035 |
| I83 | 25600 (5.1) | 0.00 | 0.01 | 0.05 | 0.11 | 0.0003 |
| J30 | 52006 (10.4) | 0.00 | 0.02 | 0.04 | 0.09 | 0.0043 |
| J44 | 28185 (5.6) | 0.01 | 0.05 | 0.09 | 0.16 | 0.0104 |
| J45 | 74385 (14.8) | 0.02 | 0.05 | 0.10 | 0.15 | 0.0014 |
| K21 | 82285 (16.4) | 0.02 | 0.04 | 0.07 | 0.12 | 0.0003 |
| K44 | 61861 (12.3) | 0.02 | 0.04 | 0.07 | 0.12 | 0.0042 |
| K57 | 74399 (14.8) | 0.01 | 0.03 | 0.05 | 0.10 | 0.0003 |
| K80 | 35894 (7.1) | 0.01 | 0.03 | 0.06 | 0.10 | 0.0011 |
| M17 | 48361 (9.6) | 0.01 | 0.04 | 0.06 | 0.09 | 0.0014 |
| M19 | 92720 (18.5) | 0.01 | 0.03 | 0.05 | 0.09 | 0.0032 |

**Supplementary Table 5: Overview of quintiles of medication uptake.**

For our application of StratGWAS to medication burden, we stratified cases into quintiles, i.e., using the $20\%-, 40\%-, 60\%-$, and 80%-quantiles of medication burden among cases as cut-off points for subgroups. Here, medication burden was defined as the number of unique medications prescribed within the BNF category matching that if the disease based on ICD-10 code, normalized by age. Columns present the cut-off points for medication burden. The highest medication uptake was observed for cases of diseases relating to the circulatory system, while lowest uptake was observed for cases in the musculoskeletal system.

| | | | | Multivariate | | Univariate | |
|---|---|---|---|---|---|---|---|
| Stratification variable | Subgroup | N | $h^2_{\text{liab}}$ | Weight | SE | Weight | SE |
| ICD10 - F32 | No | 87478 | 0.06 | 0.00 | 0.00 | 0.00 | 0.02 |
| | Yes | 41142 | 0.16 | 0.05 | 0.00 | 0.47 | 0.01 |
| ICD10 - F33 | No | 125890 | 0.09 | 0.00 | 0.00 | 0.00 | 0.04 |
| | Yes | 2730 | 0.20 | -0.01 | 0.01 | -0.07 | 0.05 |
| Phenotype | Doctor only | 86461 | 0.07 | 0.00 | 0.00 | 0.00 | 0.08 |
| | Psychiatrist only | 2273 | 0.09 | -0.04 | 0.01 | -0.36 | 0.14 |
| | Both | 39036 | 0.16 | 0.05 | 0.01 | 0.52 | 0.02 |
| Sought treatment | No | 18104 | 0.06 | 0.00 | 0.01 | 0.00 | 0.04 |
| | Yes | 28671 | 0.16 | 0.05 | 0.00 | 0.71 | 0.02 |
| Bipolar disorder | No | 126876 | 0.09 | 0.00 | 0.00 | 0.00 | 0.18 |
| | Yes | 1744 | 0.43 | -0.02 | 0.01 | 0.54 | 0.11 |
| Schizophrenia | No | 127859 | 0.09 | 0.00 | 0.00 | 0.00 | 0.27 |
| | Yes | 761 | 0.54 | -0.02 | 0.02 | 0.78 | 0.15 |
| Depressed mood | Not at all | 74377 | 0.07 | 0.00 | 0.00 | 0.00 | 0.07 |
| | Several days | 36149 | 0.14 | 0.05 | 0.00 | 0.69 | 0.10 |
| | More than half days | 6398 | 0.19 | 0.03 | 0.01 | 0.64 | 0.09 |
| | Nearly every day | 5015 | 0.26 | 0.05 | 0.01 | 0.80 | 0.07 |
| Disinterest | Not at all | 83098 | 0.07 | 0.00 | 0.00 | 0.00 | 0.07 |
| | Several days | 30814 | 0.16 | 0.04 | 0.00 | 0.77 | 0.08 |
| | More than half days | 5529 | 0.17 | 0.02 | 0.01 | 0.58 | 0.07 |
| | Nearly every day | 4193 | 0.30 | 0.05 | 0.01 | 0.88 | 0.09 |
| Restlessness | Not at all | 72445 | 0.07 | 0.00 | 0.00 | 0.00 | 0.08 |
| | Several days | 40365 | 0.14 | 0.05 | 0.00 | 0.86 | 0.11 |
| | More than half days | 5532 | 0.24 | 0.04 | 0.01 | 0.87 | 0.11 |
| | Nearly every day | 4625 | 0.38 | 0.07 | 0.01 | 1.33 | 0.07 |
| Tiredness | Not at all | 42318 | 0.08 | 0.00 | 0.00 | 0.00 | 0.09 |
| | Several days | 58616 | 0.09 | 0.04 | 0.00 | 0.84 | 0.05 |
| | More than half days | 10627 | 0.16 | 0.05 | 0.00 | 1.02 | 0.07 |
| | Nearly every day | 13538 | 0.27 | 0.08 | 0.01 | 1.63 | 0.09 |

**Supplementary Table 6: Summary and number of cases in strata of major depressive disorder.** We analyzed major depressive disorder in the UK Biobank using several stratification variables, based on modes of diagnosis, comorbidity, and depression-related symptoms. Here we present the sample sizes of each subgroup, their estimated heritability on the liability scale, and StratGWAS-derived weights when analyzing the stratification variables jointly or individually.

For all stratification variables considered, subgroups showing multiple modes of diagnosis, comorbidity, or more depressive symptoms, were associated with higher heritability compared to reference group and were assigned higher weights in the StratGWAS-derived phenotype. These weights were particularly high when analyzing stratification variables individually, which likely reflects that disease subgroups are correlated and have lower marginal effects in joint analysis.

| Sample size | Number of strata | Threads | Memory (GB) | Wall time (h) | CPU hours |
|---|---|---|---|---|---|
| 100k | 5 | 1 | 6.24 | 1.33 | 1.33 |
| | | 2 | 6.27 | 1.08 | 2.16 |
| | | 5 | 6.43 | 0.94 | 4.71 |
| | | 10 | 6.64 | 0.79 | 7.86 |
| | 20 | 1 | 7.51 | 2.70 | 2.70 |
| | | 2 | 7.73 | 1.87 | 3.74 |
| | | 5 | 7.58 | 1.32 | 6.60 |
| | | 10 | 8.95 | 1.02 | 10.20 |
| | 50 | 1 | 11.82 | 6.11 | 6.11 |
| | | 2 | 11.63 | 3.63 | 7.26 |
| | | 5 | 11.37 | 2.17 | 10.85 |
| | | 10 | 11.61 | 1.63 | 16.28 |
| 367k | 5 | 1 | 19.89 | 5.27 | 5.27 |
| | | 2 | 19.90 | 4.15 | 8.30 |
| | | 5 | 20.52 | 3.70 | 18.52 |
| | | 10 | 20.99 | 3.22 | 32.19 |
| | 20 | 1 | 20.48 | 11.40 | 11.40 |
| | | 2 | 21.44 | 7.32 | 14.64 |
| | | 5 | 21.08 | 4.63 | 23.16 |
| | | 10 | 31.76 | 4.06 | 40.60 |
| | 50 | 1 | 19.73 | 21.23 | 21.23 |
| | | 2 | 19.59 | 14.61 | 29.23 |
| | | 5 | 20.08 | 7.26 | 36.29 |
| | | 10 | 32.83 | 5.62 | 56.25 |

**Supplementary Table 7: Computational demands of StratGWAS.**

We analyzed five diseases recorded in the UK Biobank based on ICD-10 codes (E03, E11, E14, E66, and E78) using StratGWAS, using age-at-onset as stratification variable. We compared the computational demands of StratGWAS when analyzing two different sample sizes (100 k and 367 k), different number of strata (5, 20, and 50), and different number of threads (1, 2, 5, 10, 20) for parallelization. Here we report the average memory used, as well as run time and CPU hours (run time $\times$ number of threads).
